# Severity of Illness Caused by Severe Acute Respiratory Syndrome Coronavirus 2 Variants of Concern in Children: A Single-Center Retrospective Cohort Study

**DOI:** 10.1101/2021.10.23.21265402

**Authors:** Priya R. Edward, Ramon Lorenzo-Redondo, Megan E. Reyna, Lacy M. Simons, Judd F. Hultquist, Ami B. Patel, Egon A. Ozer, William J. Muller, Taylor Heald-Sargent, Matthew McHugh, Taylor J. Dean, Raj M. Dalal, Jordan John, Shannon C. Manz, Larry K. Kociolek

## Abstract

**Background:** Recent surges in coronavirus 2019 disease (COVID-19) is attributed to the emergence of more transmissible severe acute respiratory syndrome coronavirus 2 (SARS-CoV-2) variants of concern (VOCs). However, the relative severity of SARS-CoV-2 VOCs in children is unknown.

**Methods:** This retrospective single-center cohort study was performed at the Ann & Robert H. Lurie Children’s Hospital of Chicago, academic free-standing children’s hospital. We included all children ≤ 18 years-old diagnosed with COVID-19 between October 15^th^, 2020 and August 31^st^, 2021 and whose SARS-CoV-2 isolate was sequenced using the Illumina platform. For each patient sample, we identified the SARS-CoV-2 lineage, which was assigned to one of the following groups: Non-VOC, alpha VOC, beta VOC, gamma VOC, or delta VOC. We measured frequency of 5 markers of COVID-19 severity: hospitalization; COVID-19 pharmacologic treatment; respiratory support; intensive care unit admission; and severe disease as classified by the COVID-19 World Health Organization (WHO) Clinical Progression Scale (severe disease; score ≥ 6). A series of logistic regression models were fitted to estimate odds of each severity marker with each VOC (in comparison to non-VOCs), adjusting for COVID-19 community incidence and demographic and clinical co-variates.

**Results:** During the study period, 2,025 patients tested positive for SARS-CoV-2; 1,422 (70.2%) had sufficient viral load to permit sequencing. Among the 499 (35.1%) patients whose isolate was sequenced, median (inter-quartile range) age was 7 (1,12) years; 256 (51.3%) isolates were a VOC: 96 (37.5%) alpha, 38 (14.8%) gamma, and 119 (46.5%) delta. After adjusting for age, Black race, Hispanic ethnicity, high-risk medical conditions, and COVID-19 community incidence, neither alpha nor delta was associated with severe COVID-19. Gamma was independently associated with hospitalization (OR 5.9, 95% CI 1.6-21.5, *p*=0.007), respiratory support (OR 8.3, 95% CI 1.5-56.3, *p*=0.02), and severe disease as classified by the WHO Clinical Progression Scale (OR 7.7, 95% CI 1.0-78.1, *p*=0.05).

**Conclusions:** Compared to non-VOC COVID-19 infections, the gamma VOC, but not the alpha or delta VOCs, was associated with increased severity. These data suggest that recent increased in pediatric COVID-19 hospitalizations are related to increased delta COVID-19 incidence rather than increased delta virulence in children.

## Introduction

Coronavirus disease 2019 (COVID-19) is generally associated with less severe outcomes in children than adults.^1^ However, COVID-19 morbidity and mortality in children is non-trivial and may escalate because of current lack of an authorized vaccine for all pediatric age groups, return to in-person learning in schools and daycare that may promote respiratory viral transmission, and the emergence of variants of concern (VOCs) that may be associated with increased transmissibility, immune evasion, and/or worse clinical outcomes. Despite high prevalence of several VOCs worldwide and data suggesting worse clinical outcomes associated with some VOCs, some reports^2^ have not yet been peer reviewed or validated. Further, little is known about VOC-specific clinical outcomes in children.^3^

Severe acute respiratory syndrome coronavirus 2 (SARS-CoV-2) genomic surveillance data reported by the Centers for Disease Control and Prevention (CDC)^4^ indicated that the alpha (B.1.1.7 and sub-lineages) and gamma (P.1 and sub-lineages) variants emerged and expanded in midwestern US during the winter and spring of 2021. This was followed by the emergence and near complete predominance of the delta variant (B.1.617.2 and sub-lineages) in the summer of 2021. Increased morbidity related to the gamma variant, particularly in younger individuals, has been reported outside the US.^5^ Further, concomitant with surges of the more transmissible delta variant, increasing pediatric COVID-19 incidence and subsequent increases in pediatric COVID-19 hospitalizations have been observed.^6, 7^ However, it is unclear if increases in hospitalizations are solely related to increased COVID-19 incidence or if certain VOCs are associated with more severe disease.

At our children’s hospital, we perform genomic surveillance of SARS-CoV-2 in children. The objective of this single-center retrospective cohort study was to compare COVID-19 severity in children with prevalent VOCs compared to those infected with other SARS-CoV-2 lineages. These data provide a framework for comparing outcomes in newly emerging variants. Further, understanding the association between VOCs and clinical outcomes is essential to guide public health response and prepare children’s hospitals for expected healthcare resource needs based on observed SARS-CoV-2 molecular epidemiology.

## Methods

### Setting and Participants

This retrospective cohort study was conducted at the Ann & Robert H. Lurie Children’s Hospital of Chicago, a tertiary care academic free-standing children’s hospital. The Lurie Children’s Institutional Review Board approved this study (IRB 2020-3792). Using residual diagnostic samples from pediatric inpatients and outpatients who tested positive by SARS-CoV-2 polymerase chain reaction (PCR) in our clinical microbiology laboratory, we perform surveillance for SARS-CoV-2 variants using whole genome sequencing (WGS) with the Center for Pathogen Genomics and Microbial Evolution at Northwestern University. As part of our ongoing surveillance efforts, samples were eligible for WGS if they had a sufficient quantity of residual viral transport media remaining after clinical COVID-19 testing and a sufficiently low PCR cycle threshold for the SARS-CoV-2 N open reading frame (i.e., sufficiently high viral load). Twice monthly, approximately 25-50% of samples meeting these criteria are randomly selected for WGS. For the present study, all patients ≤ 18 years-old at our children’s hospital with sequenced SARS-CoV-2 isolates between October 15^th^, 2020 and August 31^st^, 2021 were included. If patients had more than one positive sample, only the first sample and associated clinical encounter was included.

### SARS-CoV-2 Whole Genome Sequencing

Laboratory methods for viral RNA extraction, cDNA synthesis, viral genome amplification, sequencing library preparation, Illumina sequencing, genome assembly, and bioinformatics analyses are described in the Supplemental Materials. Consensus genome sequences were deposited in the GISAID public database (Table S1).

### Study Data and Definitions

Manual chart review was performed for all subjects. The following demographic and clinical information were collected: age, sex, race, ethnicity, presence of COVID-19 symptoms, respiratory viral co-infections, COVID-19 vaccine status, receipt of SARS-CoV-2 monoclonal antibodies for mild early infection in high-risk patients, and co-morbidities. Presence of COVID-19 symptoms was defined as any of the following listed in the medical record at the time of COVID-19 testing: fever, rhinorrhea, congestion, sore throat, cough, shortness of breath, loss of taste or smell, headache, or abdominal pain. Respiratory viral co-infections were identified by PCR within 7 days of COVID-19 diagnosis. Children were further classified as high-risk for COVID-19 complications if they had an underlying condition associated with this higher risk per high-quality and reproducible data (i.e., meta-analysis, systematic review, or observational study; not small studies, case reports/series, or conflicting evidence) based on CDC classification as of August 31^st^, 2021 (Table S2).^8^ Children were considered fully vaccinated if at least two weeks had elapsed between completing the vaccination series and onset of their COVID-19 illness.

Charts were reviewed for several markers of COVID-19 severity, including hospitalization for COVID-19; pharmacologic treatment for COVID-19 (i.e., including remdesivir, corticosteroids, or tocilizumab, but excluding monoclonal antibodies because these are given to high-risk outpatients early in infection to prevent subsequent morbidity); respiratory support; intensive care unit (ICU) admission for COVID-19; and severe disease as classified by the COVID-19 World Health Organization (WHO) Clinical Progression Scale^9^ (score ≥ 6; Table S3); and death caused by COVID-19. Based on the lineage of SARS-CoV-2 identified by WGS, children were grouped based on whether their COVID-19 infection was caused by a VOC (i.e., alpha, beta, gamma, or delta), as well as the specific VOC lineage, using CDC classification as of August 31^st^, 2021.^2^ Of note, on September 21^st^, 2021 after completion of this study, the alpha, beta, and gamma VOCs were reclassified as “variants being monitored” by the CDC because US incidence subsequently declined in a significant and sustained manner.

### Statistical Analyses

Demographic, clinical, and outcome categorical variables for the entire cohort, as well as for patient subgroups infected with either a non-VOC, the alpha VOC, the gamma VOC, or the delta VOC, were summarized and reported as frequencies and percentages. Frequencies of these variables among subgroups were compared using Pearson’s chi-squared test, and continuous variables were compared using Kruskal-Wallis test. Next, to assess the clinical severity associated with each VOC, a series of logistic regression models were fitted to estimate odds of each severity marker of interest with each VOC in comparison to non-VOCs, adjusting for COVID-19 incidence and relevant demographic and clinical co-variates associated with COVID-19 outcomes, including Black race,^10^ Hispanic ethnicity,^10^ age,^1^ and high-risk medical conditions.^8^ We also adjusted for trends in pediatric COVID-19 community incidence using publicly available data from the Chicago Department of Public Health. Odds ratios (OR) and 95% confidence intervals (CI) were estimated and tested for statistical significance for each VOC included in the model compared to the non-VOC group. Two-sided *P* values < 0.05 were considered statistically significant. Statistical analyses were performed using Stata/IC 16.0 (StataCorp, College Station, TX) and R version 4.1.0.

## Results

During the 10-month study period (Oct. 15^th^, 2020 -Aug. 31^st^, 2021), 2,025 patients tested positive for SARS-CoV-2 by PCR. Of these, 1,622 (80.1%) had symptoms of COVID-19. The remaining 403 (19.9%) were asymptomatic and tested for screening at the time of admission (n=74), prior to a surgical procedure (n=258), or other reasons (n=71) such as post-exposure testing or pre/post-travel screening testing. PCR cycle threshold values were too high to permit successful sequencing in 603 (29.8%) samples. Of the 1,422 patients with samples with sufficient viral load, samples from 499 (35.1%) patients 0-18 years underwent WGS, and these patients were included in the study. Median (IQR) age of patients included in this study was 7 (1,12) years-old; age range was <1 month - 18 years old. Of the 499 sequenced isolates, 256 (51.3%) were classified a VOC, including 96 (37.5%) alpha, 3 (1.2%) beta, 38 (14.8%) gamma, and 119 (46.5%) delta. Because of the very low prevalence of beta, these 3 patients were excluded from further analyses.

Figure 1 illustrates frequencies (Figure 1A) proportions (Figure 1B) of SARS-CoV-2 lineages identified during the study period. The specific isolate lineages included are listed in Table S1. Alpha and gamma emerged in January and February 2021, respectively. Together, the alpha and gamma VOCs accounted for most pediatric COVID-19 infections by May 2021 (Fig 1A), at which time COVID-19 incidence in children was declining as evidenced both by number of pediatric samples available for sequencing (Figure 1B) and Chicago COVID-19 incidence (Figure 2). COVID-19 incidence reached the pandemic nadir in Chicago in adults and children in June 2021, shortly after which incidence increased dramatically with the emergence of delta. Our trends in variant epidemiology generally resembled that observed in adults in Chicago during the same period (Figure S1).

**Figure 1:**
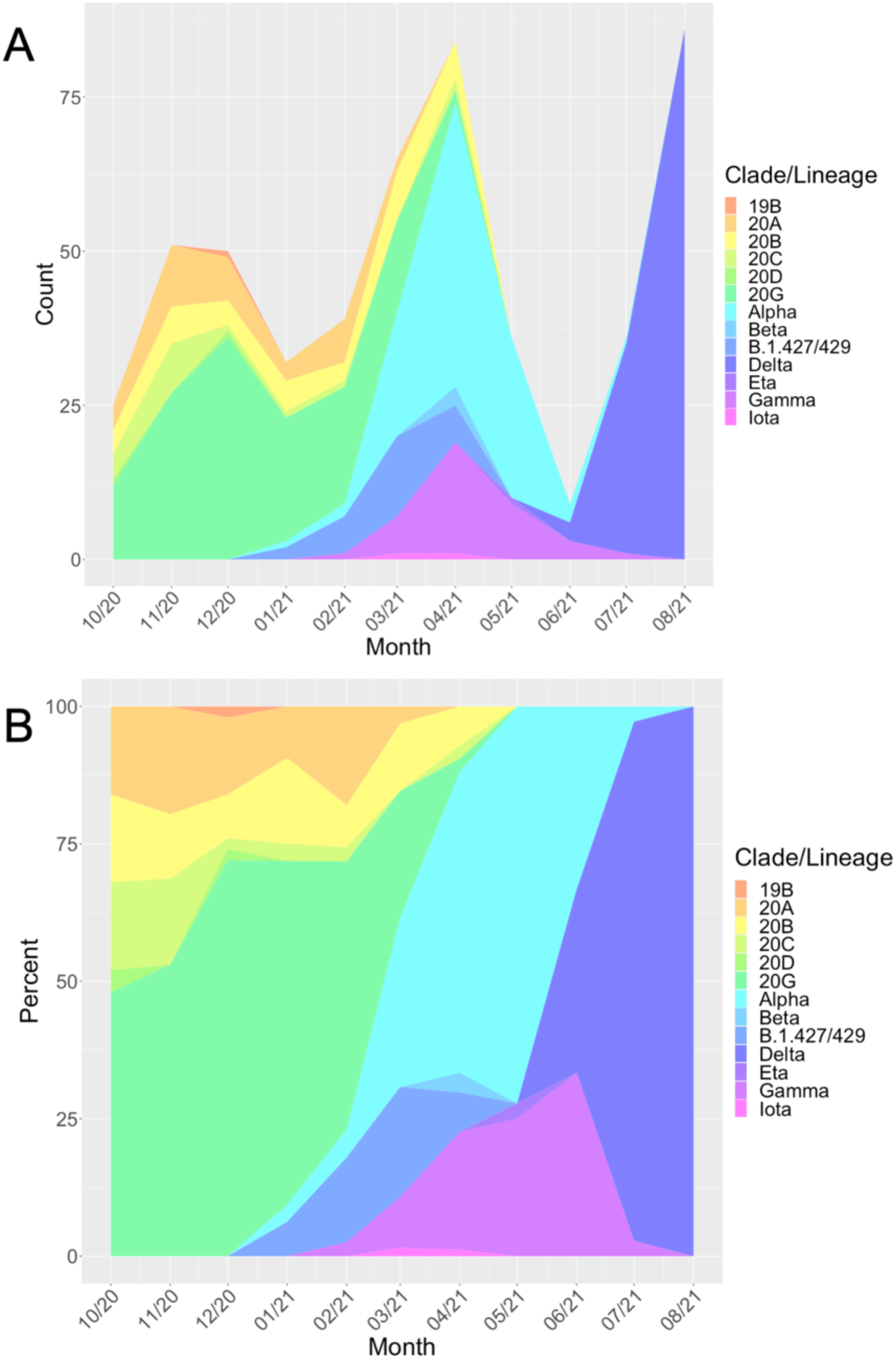
SARS-CoV-2 Molecular and Clinical Epidemiology. Frequencies (A) and proportions (B) of lineages of SARS-CoV-2 identified in pediatric patients during 10-month study period (October 2020 – August 2021). Variants of concern identified include alpha, beta, gamma, and delta. Corresponding data for adult patients at a partner hospital in Chicago are shown in Figure S1; the variant proportions and chronology in adults generally resembled what was observed in children during the same period.

**Figure 2:**
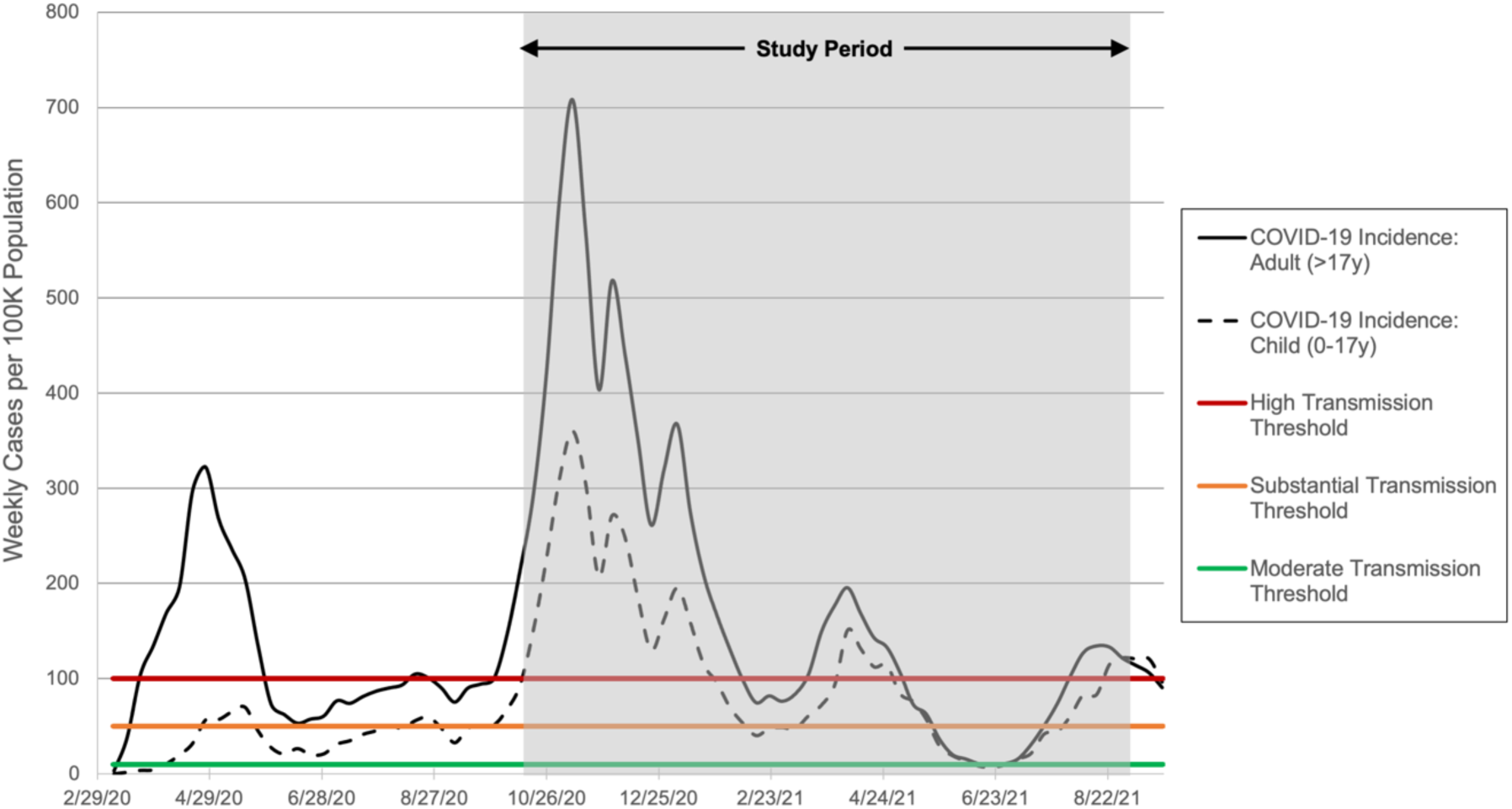
COVID-19 Incidence in Chicago Children and Adults. Incidence of COVID-19 (weekly cases per 100K population) in children (age 0-17 years [y]) and adults (age >17y) in Chicago throughout the pandemic and during the study period (shaded) during which the sequencing data presented here were derived. Thresholds for community transmission levels as defined by the CDC^20^ are noted with colored vertical lines.

Clinical, demographic, and outcome data for the entire cohort, and for subgroups of patients infected with a non-VOC, alpha VOC, gamma VOC, or delta VOC, are listed in Table 1. Statistically significant differences between these subgroups were observed in race (greater proportion of patients who identify as Black in those infected with VOCs) and in proportion of patients who received outpatient treatment with casirivimab-imdevimab for early mild COVID-19 (all three patients were infected with alpha). Two children were fully vaccinated at the time of developing COVID-19, one each infected with gamma and delta, but proportions did not significantly differ between groups. Statistically significant differences between subgroups were also observed for three markers of severity – hospitalization for COVID-19, respiratory support, and severe disease as classified by the WHO Clinical Progression Scale (score ≥6; Table S3). In each case, frequency of these outcomes was highest in those children infected with gamma. There was one COVID-19-associated death, and this was associated with a non-VOC infection.

**Table 1:** Demographics, clinical characteristics, and markers of COVID-19 severity of 499 children with COVID-19.

|  | Entire Cohort,<br>n (%)<br>(n=499) | Non-VOC,<br>n (%)<br>(n=243) | Alpha,<br>n (%)<br>(n=96) | Gamma,<br>n (%)<br>(n=38) | Delta,<br>n (%)<br>(n=119) | p value |
| --- | --- | --- | --- | --- | --- | --- |
| Age (years; median [IQR]) | 7 (1,12) | 7 (1,12) | 9 (2,14) | 7 (1,14) | 4 (1,11) | 0.09 |
| 12-18y | 147 (29.4%) | 75 (30.8%) | 38 (39.5%) | 13 (34.2%) | 21 (17.6%) |  |
| 6-11y | 120 (24.0%) | 54 (22.2%) | 21 (21.8%) | 9 (23.6%) | 33 (27.7%) |  |
| 1-5y | 144 (28.8%) | 71 (29.2%) | 22 (22.9%) | 8 (21.0%) | 43 (36.1%) |  |
| 0-12m | 88 (17.6%) | 43 (17.6%) | 15 (15.6%) | 8 (21.0%) | 22 (18.4%) |  |
| Sex (male) | 246 (49.2%) | 124 (51.0%) | 47 (48.9%) | 21 (55.2%) | 53 (44.5%) | 0.59 |
| Race (Black) | 79 (15.8%) | 22 (9.0%) | 24 (25%) | 8 (21.0%) | 25 (21.0%) | **0.001\*** |
| Ethnicity (Hispanic) | 212 (42.4%) | 117 (48.1%) | 37 (38.5%) | 16 (42.1%) | 42 (35.2%) | 0.098 |
| High-risk condition for COVID-19 complications | 167 (33.5%) | 81 (33.3%) | 40 (41.7%) | 13 (34.2%) | 33 (27.7%) | 0.20 |
| Respiratory viral co-infection | 1 (0.2%)^ | 0 (0%) | 0 (0%) | 0 (0%) | 1 (0.8%)^ | 0.37 |
| Fully vaccinated against COVID-19 | 2 (0.4%) | 0 (0%) | 0 (0%) | 1 (2.6%) | 1 (0.8%) | 0.25 |
| Outpatient treatment with casirivimab-imdevimab | 3 (0.6%) | 0 (0%) | 3 (3.1%) | 0 (0%) | 0 (0%) | **0.006\*** |
| Symptoms of COVID-19 | 455 (91.1%) | 223 (91.7%) | 88 (91.6%) | 34 (89.4%) | 107 (89.9%) | 0.92 |
| Hospitalized for COVID-19 | 29 (5.8%) | 8 (3.2%) | 7 (7.2%) | 6 (15.7%) | 8 (6.7%) | **0.017\*** |
| COVID-19 pharmacologic treatment | 13 (2.6%) | 4 (1.6%) | 2 (2.0%) | 3 (7.8%) | 4 (3.3%) | 0.14 |
| *remdesivir* | 12 (2.4%) | 4 (1.6%) | 1 (1.0%) | 3 (7.8%) | 4 (3.3%) |  |
| *dexamethasone* | 9 (1.8%) | 3 (1.2%) | 1 (1.0%) | 3 (7.8%) | 2 (1.6%) |  |
| *tocilizumab* | 1 (0.2%) | 1 (0.4%) | 0 (0%) | 0 (0%) | 0 (0%) |  |
| Respiratory support | 10 (2.0%) | 3 (1.2%) | 0 (0%) | 4 (10.5%) | 3 (2.5%) | **0.001\*** |
| *Nasal cannula* | 3 (0.6%) | 1 (0.4%) | 0 (0%) | 1 (2.6%) | 1 (0.8%) |  |
| *High-flow nasal cannula* | 1 (0.2%) | 0 (0%) | 0 (0%) | 1 (2.6%) | 0 (0%) |  |
| *Continuous positive airway pressure* | 0 (0%) | 0 (0%) | 0 (0%) | 0 (0%) | 0 (0%) |  |
| *Bilevel positive airway pressure* | 4 (0.8%) | 1 (0.4%) | 0 (0%) | 2 (5.2%) | 1 (0.8%) |  |
| *Mechanical ventilation* | 2 (0.4%) | 1 (0.4%) | 0 (0%) | 0 (0%) | 1 (0.8%) |  |
| Intensive care unit admission | 9 (1.8%) | 3 (1.2%) | 1 (1.0%) | 2 (5.2%) | 3 (2.5%) | 0.30 |
| WHO Clinical Progression Scale $\geq 6$ | 7 (1.4%) | 2 (0.8%) | 0 (0%) | 3 (7.8%) | 2 (1.6%) | **0.004\*** |
| *0-2 (mild)* | 461 (92.3%) | 230 (94.6%) | 85 (88.5%) | 32 (84.2%) | 111 (93.2%) |  |
| *3-5 (moderate)* | 31 (6.2%) | 11 (4.5%) | 11 (11.4%) | 3 (7.8%) | 6 (5.0%) |  |
| *6-9 (severe)* | 6 (1.2%) | 1 (0.4%) | 0 (0%) | 3 (7.8%) | 2 (1.6%) |  |
| *10 (morbid)* | 1 (0.2%) | 1 (0.4%) | 0 (0%) | 0 (0%) | 0 (0%) |  |
| COVID-19-related death | 1 (0.2%) | 1 (0.4%) | 0 (0%) | 0 (0%) | 0 (0%) | 0.79 |

IQR- interquartile range. \*Bolded values indicate statistical significance ( $p < 0.05$ ). ^Co-infection with rhinovirus/enterovirus.

To determine the association between each VOC (in comparison to non-VOC infections) and clinical severity, we fitted a series of logistic regression models for five markers of disease severity, adjusting for temporal changes in COVID-19 incidence and relevant clinical and demographic characteristics (Table 2). For COVID-19 pharmacologic treatment, respiratory support, ICU admission, and severe disease as classified by the WHO Clinical Progression Scale, increasing age was associated with these severity markers, with odds increasing by ∼10-20% for each year increase. High-risk conditions were independently associated with hospitalization (OR 6.1, 95% CI 2.6-15.9, *p*<0.001); odds of the other severity markers could not be calculated because high-risk conditions were mutually inclusive with those severity markers. Compared to non-VOCs, infection with alpha or delta was not associated with any markers of COVID-19 severity. However, infection with gamma was independently associated with hospitalization (OR 5.9, 95% CI 1.6-21.5, *p*=0.007), respiratory support (OR 8.3, 95% CI 1.5-56.3, *p*=0.02), and severe disease as classified by the WHO Clinical Progression Scale (OR 7.7, 95% CI 1.0-78.1, *p*=0.05).

**Table 2:**
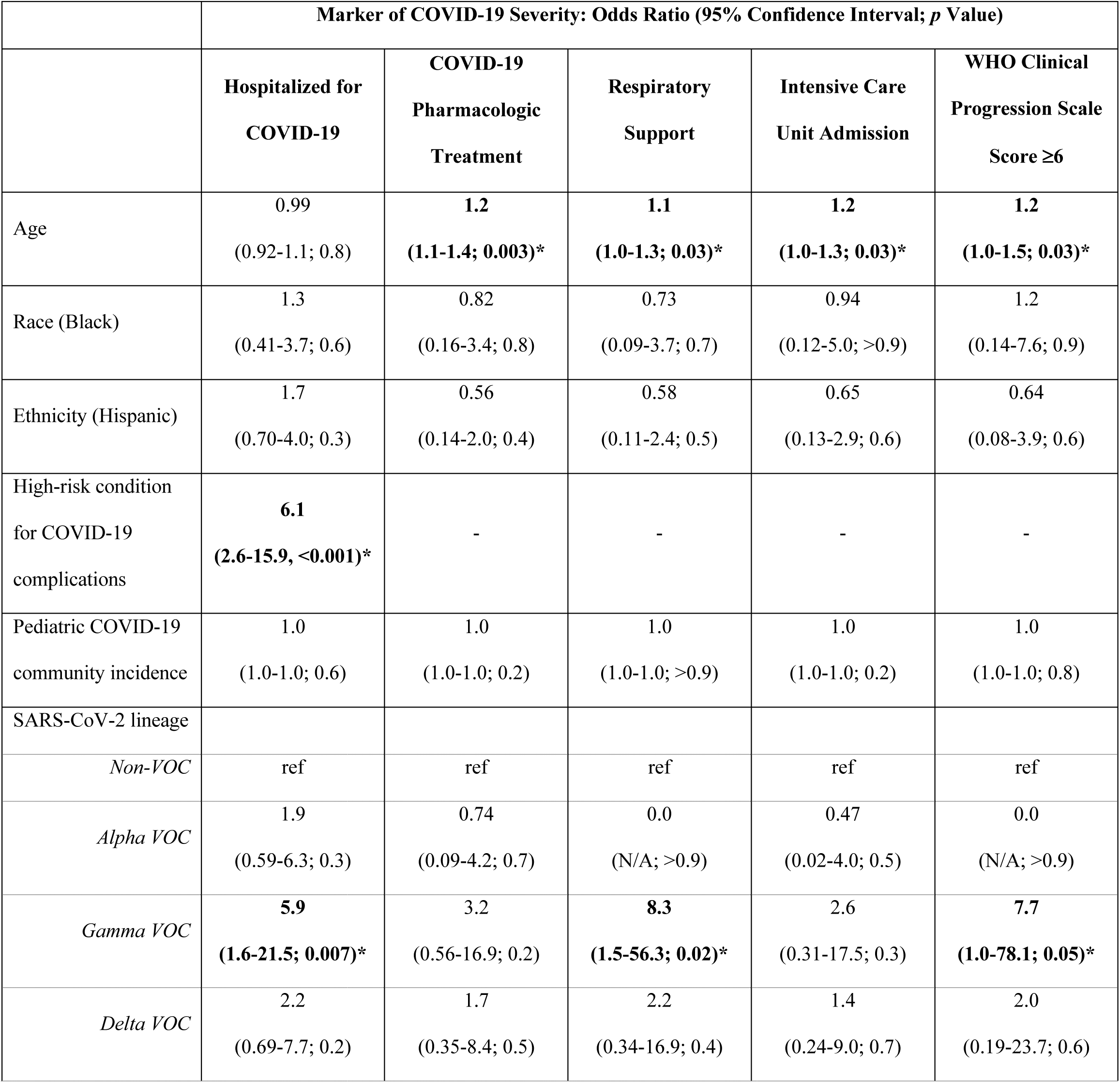
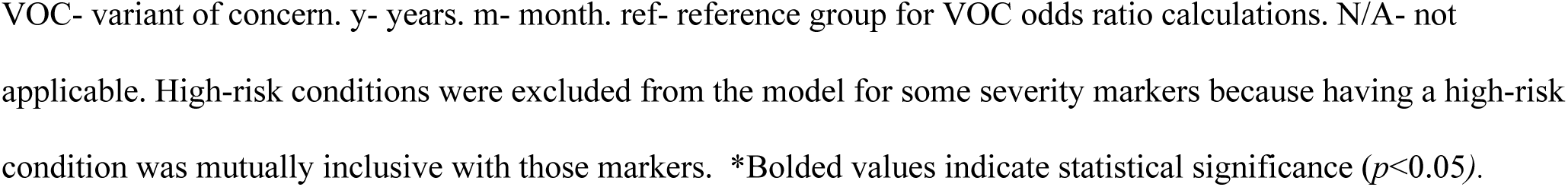
Association between COVID-19 severity and SARS-CoV-2 variants of concern.

A small number of patients were fully vaccinated (n=2; one each in the gamma and delta VOC groups) at time of their breakthrough infection or had received SARS-CoV-2 monoclonal antibodies (n=3; all in the alpha VOC group) as an outpatient early in infection to prevent subsequent morbidity. Because vaccines and monoclonal antibodies are given to reduce COVID-19-associated complications, a sensitivity analysis was performed whereby we repeated the models presuming that all five patients would have developed each marker of severity had they not received these therapies (Table S4). There were no significant changes in association between VOCs and clinical outcomes following this sensitivity analysis.

## Discussion

In this single center retrospective cohort study of children with COVID-19 in the Chicago area, severe COVID-19 outcomes were relatively rare, confirming previously findings of COVID-19 in children.^1^ SARS-CoV-2 infection with gamma (P.1 and sub-lineages), but not alpha (B.1.1.7 and sublineages) or delta (B.1.617.2 and sub-lineages), was associated with increased COVID-19 severity as evidenced by its association with hospitalization for COVID-19, respiratory support, and WHO Clinical Progression Scale score ≥ 6. These findings support previous observations in Brazil^6^ whereby younger patients without comorbidities were hospitalized with severe COVID-19 infections during a surge in gamma SARS-CoV-2 infections between December 2020 and January 2021. These findings also support a prior US study that demonstrated significantly increased number of COVID-19 pediatric hospitalizations during nationwide surges with the more transmissible delta VOC, but the overall proportion of hospitalizations requiring the ICU or mechanical ventilation did not change following delta emergence.^7^ However, our findings in children differ from that in adults in Canada where N501Y VOCs (i.e., alpha, beta, and gamma collectively) and delta was associated with increased COVID-19 morbidity and mortality.^11^

During the study period, alpha and gamma expanded and caused the vast majority of pediatric infections in late winter and early spring of 2021. However, we identified expansion of the proportion of alpha and gamma VOCs in our population concomitant with significant declines in pediatric and adult COVID-19 incidence in the Chicago area. This suggests that despite increased severity associated with gamma in children, gamma was unable to sustain high-level transmission, even in a largely unvaccinated pediatric population where many, but not all, schools in the area had returned to the classroom, albeit with risk mitigation measures in place. In summer of 2021, delta expanded throughout the US and has completely replaced the alpha and gamma VOCs. The reasons delta has replaced alpha and gamma are still unclear but likely related to the higher transmissibility of the delta variant.^12, 13^

More severe outcomes in children associated with specific VOCs suggest value in developing assays that rapidly distinguish circulating variants. These findings also suggest that targeted interventions in areas where gamma is circulating should be considered, such as implementing a lower threshold for monoclonal antibody administration, and avoidance of empiric use of monoclonal antibodies to which the gamma VOC is resistant (e.g., bamlanivimab and etesevimab).^14^ Clinical risk management in children could be facilitated based on suspicion of gamma, either based on rapid in-patient genotyping or community surveillance. Perhaps reassuringly, as of October 2021,^15^ gamma VOC incidence has declined substantially worldwide, causing more than 1% of new infections only in Peru (10.1%), Brazil (6.1%), and Chile (4.3%). Similar incidence trends have been identified for alpha. For these reasons, alpha and gamma have recently been reclassified as “variants being monitored,” rather than VOCs, by the CDC.

The reasons for the observed association between gamma and COVID-19 severity in children in this study remain hypothetical. Gamma shares several mutations in the receptor-binding domain (RBD) of the spike protein with other VOCs, including several that have been previously linked with humoral immune evasion in adults (i.e., K417T, E484K, and N501Y).^16^ There are similarly several mutations unique to the gamma VOC in the N-terminal domain of spike (L18F, T20N, P26S, D138Y, R190S). While many of these mutations map to regions of neutralizing antibody binding,^17^ the impact of these mutations on neutralization remains unclear. More research on the neutralizing antibody responses to SARS-CoV-2 in children and their relationship to these mutations is needed. While differential susceptibility to antibody neutralization may be one explanation, several VOCs besides gamma would be expected to confer this phenotype.^17^ An alternative explanation is a difference in cellular tropism unique to gamma in children. Gamma carries a unique mutation in the membrane-proximal heptad repeat region (HR2) of Spike responsible for fusion activity and tropism, V1176F.^18^ It is shared by the Zeta (P.2) and Theta (P.3) variants; further analysis of disease outcome associations with these variants may be informative. Finally, gamma carries several other mutations in other open reading frames (ORFs) that may contribute to its pathogenesis (i.e., S1188L, K1795Q, del3675-3677 in ORF1a; P314L, E1264D in ORF1b; S253P in ORF3a; S84L, E92K in ORF8; and P80R, R203K, G204R in N),^19^ though more research on the functional impact of these mutations on the viral lifecycle and host response is needed.

A notable strength of this study includes classification of patients based on confirmed SARS-CoV-2 lineage, which limited misclassification bias; prior studies ^6, 7^ compared patient outcomes during different time periods, which was used as a proxy of SARS-CoV-2 lineage. Use of monoclonal antibodies and breakthrough infections following COVID-19 vaccination were rare events in this study. Nonetheless, we performed a sensitivity analysis whereby we repeated the models presuming that these five patients would have developed each marker of severity had they not received these therapies. Our findings were consistent, suggesting limited bias from the protective impact of monoclonal antibodies and vaccines.

This study has several limitations. As a single-center study, our findings may not be generalizable to other geographic locations and patient populations. However, the referral area for our hospital in Chicago extends throughout the city and suburban areas and includes a diverse patient base in terms of demographics and medical complexity. These findings should be confirmed in other pediatric and adult populations. Further, the relatively small sample size and rarity of severe outcomes in children contributed to wide CIs, reducing the precision of the effect estimates for the association between the gamma VOC and severe COVID-19 outcomes. This study was limited to those infections for which SARS-CoV-2 was able to be sequenced, biasing the study to inclusion of infections associated with relatively low cycle thresholds (i.e., high viral loads). Although we performed multivariable analyses, it is possible that unmeasured confounding has contributed to our findings. Although we limited our definition of high risk for COVID-19 complications to those for which there are high-quality and reproducible data (i.e., meta-analysis, systematic review, or observational study; not small studies, case reports/series, or conflicting evidence) per CDC classification. (Table S2),^8^ these data are predominantly from adult populations and may not all apply to children. Nonetheless, based on our observations of COVID-19 severity being strongly associated with, and sometimes mutually inclusive with, having a high-risk condition, those adult data likely translate well to children.

In summary, compared to non-VOC COVID-19 infections, the gamma VOC, but not the alpha or delta VOCs, was associated with a more severe outcome in our single-center pediatric cohort. These data suggest that recent increases in pediatric COVID-19 hospitalizations are related to increased delta COVID-19 incidence rather than increased delta virulence in children. Ongoing genomic surveillance should prioritize identifying re-emergence of the gamma variant in the US, as well as identifying emergence and clinical outcomes associated with future variants. Investigation of benefits of more aggressive pharmacologic interventions may be warranted in the few areas with ongoing gamma transmission. Although children generally have mild COVID-19 infection, the emergence of more virulent VOCs could dramatically change our clinical and public health response to COVID-19 in children through our iterative risk-benefit analyses of public health and pharmacologic interventions.

## Data Availability

All data produced in the present work are contained in the manuscript

## Acknowledgements

The authors acknowledge the staff in the Clinical Microbiology and Special Infectious Diseases laboratories at Lurie Children’s for their assistance in saving and processing specimens for WGS. We also acknowledge Katherine Doyle from the Chicago Department of Public Health for providing the Chicago SARS-CoV-2 incidence data by age group. This research was supported in part through the computational resources and staff contributions provided for the Quest high performance computing facility at Northwestern University, which is jointly supported by the Office of the Provost, the Office for Research, and Northwestern University Information Technology.

## References

1. Coronavirus Disease 2019 in Children - United States, February 12-April 2, 2020. MMWR Morbidity and mortality weekly report. Apr 10 2020;69(14):422–426. doi:10.15585/mmwr.mm6914e4

2. Centers for Disease Control and Prevention; SARS-CoV-2 Variant Classifications and Definitions. Accessed August 31st, 2021, https://www.cdc.gov/coronavirus/2019-ncov/variants/variant-info.html

3. Brookman S, Cook J, Zucherman M, Broughton S, Harman K, Gupta A. Effect of the new SARS-CoV-2 variant B.1.1.7 on children and young people. Lancet Child Adolesc Health. Apr 2021;5(4):e9–e10. doi:10.1016/s2352-4642(21)00030-4

4. Centers for Disease Control and Prevention; COVID Data Tracker: Monitoring Variant Proportions. Accessed October 13th, 2021, https://covid.cdc.gov/covid-data-tracker/#variant-proportions

5. Nonaka CKV, Gräf T, Barcia CAL, et al. SARS-CoV-2 variant of concern P.1 (Gamma) infection in young and middle-aged patients admitted to the intensive care units of a single hospital in Salvador, Northeast Brazil, February 2021. Int J Infect Dis. Oct 2021;111:47–54. doi:10.1016/j.ijid.2021.08.003

6. Delahoy MJ, Ujamaa D, Whitaker M, et al. Hospitalizations Associated with COVID-19 Among Children and Adolescents - COVID-NET, 14 States, March 1, 2020-August 14, 2021. MMWR Morbidity and mortality weekly report. Sep 10 2021;70(36):1255–1260. doi:10.15585/mmwr.mm7036e2

7. Siegel DA, Reses HE, Cool AJ, et al. Trends in COVID-19 Cases, Emergency Department Visits, and Hospital Admissions Among Children and Adolescents Aged 0-17 Years - United States, August 2020-August 2021. MMWR Morbidity and mortality weekly report. Sep 10 2021;70(36):1249–1254. doi:10.15585/mmwr.mm7036e1

8. Centers for Disease Control and Prevention; Science Brief: Evidence Used to Update the List of Underlying Medical Conditions Associated with Higher Risk for Severe COVID-19. Accessed August 31st, 2021, https://www.cdc.gov/coronavirus/2019-ncov/science/science-briefs/underlying-evidence-table.html

9. WHO Working Group on the Clinical Characterisation and Management of COVID-19 infection; A minimal common outcome measure set for COVID-19 clinical research. Lancet Infect Dis. Aug 2020;20(8):e192–e197. doi:10.1016/s1473-3099(20)30483-7

10. Centers for Disease Control and Prevention; Risk for COVID-19 Infection, Hospitalization, and Death By Race/Ethnicity. Accessed September 13th, 2021, https://www.cdc.gov/coronavirus/2019-ncov/covid-data/investigations-discovery/hospitalization-death-by-race-ethnicity.html

11. Fisman DN, Tuite AR. Evaluation of the relative virulence of novel SARS-CoV-2 variants: a retrospective cohort study in Ontario, Canada. Cmaj. Oct 4 2021;doi:10.1503/cmaj.211248

12. Public Health England; Increased household transmission of COVID-19 cases associated with SARS-CoV-2 Variant of Concern B.1.617.2: a national case control study. Accessed October 13th, 2021, https://khub.net/documents/135939561/405676950/Increased+Household+Transmission+of+COVID-19+Cases+-+national+case+study.pdf/7f7764fb-ecb0-da31-77b3-b1a8ef7be9aa

13. Earnest R, Uddin R, Matluk N, et al. Comparative transmissibility of SARS-CoV-2 variants Delta and Alpha in New England, USA. medRxiv. Oct 7 2021;doi:10.1101/2021.10.06.21264641

14. Infectious Diseases Society of America; IDSA Guidelines on the Treatment and Management of Patients with COVID-19. Accessed September 10th, 2021, https://www.idsociety.org/practice-guideline/covid-19-guideline-treatment-and-management/

15. GISAID, Most recent reported occurrences in different countries. Accessed October 14th, 2021, https://www.gisaid.org/hcov19-variants/

16. Xie X, Liu Y, Liu J, et al. Neutralization of SARS-CoV-2 spike 69/70 deletion, E484K and N501Y variants by BNT162b2 vaccine-elicited sera. Nat Med. Apr 2021;27(4):620–621. doi:10.1038/s41591-021-01270-4

17. Gobeil SM, Janowska K, McDowell S, et al. Effect of natural mutations of SARS-CoV-2 on spike structure, conformation, and antigenicity. Science (New York, NY). Aug 6 2021;373(6555)doi:10.1126/science.abi6226

18. Liu S, Xiao G, Chen Y, et al. Interaction between heptad repeat 1 and 2 regions in spike protein of SARS-associated coronavirus: implications for virus fusogenic mechanism and identification of fusion inhibitors. Lancet. Mar 20 2004;363(9413):938–47. doi:10.1016/s0140-6736(04)15788-7

19. P.1 Lineage Report. Accessed October 19th, 2021, https://outbreak.info/situation-reports?pango=P.1

20. Centers for Disease Control and Prevention; COVID Data Tracker. Accessed September 12th, 2021, https://covid.cdc.gov/covid-data-tracker/#datatracker-home

