## Supplemental Materials for "Severity of Illness Caused by Severe Acute Respiratory Syndrome Coronavirus 2 Variants of Concern in Children: A Single-Center Retrospective Cohort Study"

### Supplemental Methods

*SARS-CoV-2 Whole Genome Sequencing*

Viral RNA Extraction

Viral RNA was extracted from clinical specimens utilizing the QIAamp 96 Virus QIAcube HT Kit (Qiagen, cat. no. 57731). Clinical testing for SARS-CoV-2 presence was performed by quantitative reverse transcription and PCR (qRT-PCR) with the CDC 2019-nCoV RT-PCR Diagnostic Panel utilizing the N1 probe in SARS-CoV-2 and RP probes for sample quality control as previously described (IDT, cat. no. 10006713). All specimens that failed to amplify the RP housekeeping gene were excluded from this study. All specimens with an N1 probe cycle threshold (Ct) less than or equal to 35 were considered positive and included in this study. RT-PCR was performed on all specimens to validate Ct values obtained by the clinical diagnostic laboratory. Ct values from the N1 probes were used in all subsequent analyses.

cDNA Synthesis and Viral Genome Amplification

cDNA synthesis was performed with SuperScript IV First Strand Synthesis Kit (ThermoFisher, cat. no. 18091050) using 11 µl of extracted viral nucleic acids and random hexamers according to manufacturer's specifications. Direct amplification of the viral genome cDNA was performed in multiplexed PCR reactions to generate ~400 bp amplicons tiled across the genome. The multiplex primer set, comprised of two non-overlapping primer pools, was created using Primal Scheme and provided by the Artic Network (version 3 and 4 releases). PCR amplification was carried out using Q5 Hot Start HF Taq Polymerase (NEB, cat. no. M0493L) with 5 µl of cDNA in a 25 µl reaction volume. A two-step PCR program was used with an initial step of 98 °C for 30 s, then 35 cycles of 98 °C for 15 s followed by five minutes at 64 °C. Separate reactions were carried out for each primer pool and validated by agarose gel electrophoresis alongside negative controls. Each reaction set included positive and negative amplification controls and was performed in a space physically separated for pre- and post-PCR processing steps to reduce contamination. Amplicon sets for each genome were pooled prior to sequencing library preparation.

Sequencing Library Preparation, Illumina Sequencing, and Genome Assembly

Sequencing library preparation of genome amplicon pools was performed using the SeqWell plexWell 384 kit per manufacturer's instructions. Pooled libraries of up to 96 genomes were sequenced on the Illumina MiSeq using the V2 500 cycle kit. Sequencing reads were trimmed to remove adapters and low-quality sequences using Trimmomatic v0.36. Trimmed reads were aligned to the reference genome sequence of SARS-CoV-2 (accession MN908947.3) using bwa v0.7.15. Pileups were generated from the alignment using samtools v1.9 and consensus sequence determined using iVar v1.2.2 with a minimum depth of 10, a minimum base quality score of 20, and a consensus frequency threshold of 0 (i.e. majority base as the consensus).[1] Consensus genome sequences were deposited in the GISAID public database (Table S1).

Bioinformatics Analyses

Genome sequences were aligned using MAFFT v7.453 software and manually edited using MEGA v6.06. We used the Pango classification scheme (https://cov-lineages.org/; PangoLearn version 9/28/2021) to identify the lineages that subsequently were grouped in variants using scorpio (scorpio v0.3.12). To confirm the clustering of the sequences by lineage, we inferred a Maximum Likelihood phylogeny with IQ-Tree v2.0.5 using its ModelFinder function before each analysis to estimate the nucleotide substitution model best-fitted for each dataset by means of Bayesian information criterion (BIC). We assessed the support of the clusters formed by the different lineages both with the Shimodaira–Hasegawa approximate likelihood-ratio test (SH-aLRT) and with ultrafast bootstrap (UFboot) with 1000 replicates each.

**Supplemental Figure 1**: Frequencies (A) and proportions (B) of lineages of SARS-CoV-2 identified in adult patients during 10-month study period (October 2020 – August 2021) in a Chicago partner hospital. Corresponding data for children in this study are shown in Figure 1; the variant proportions and chronology in adults generally resembled what was observed in children during the same period.

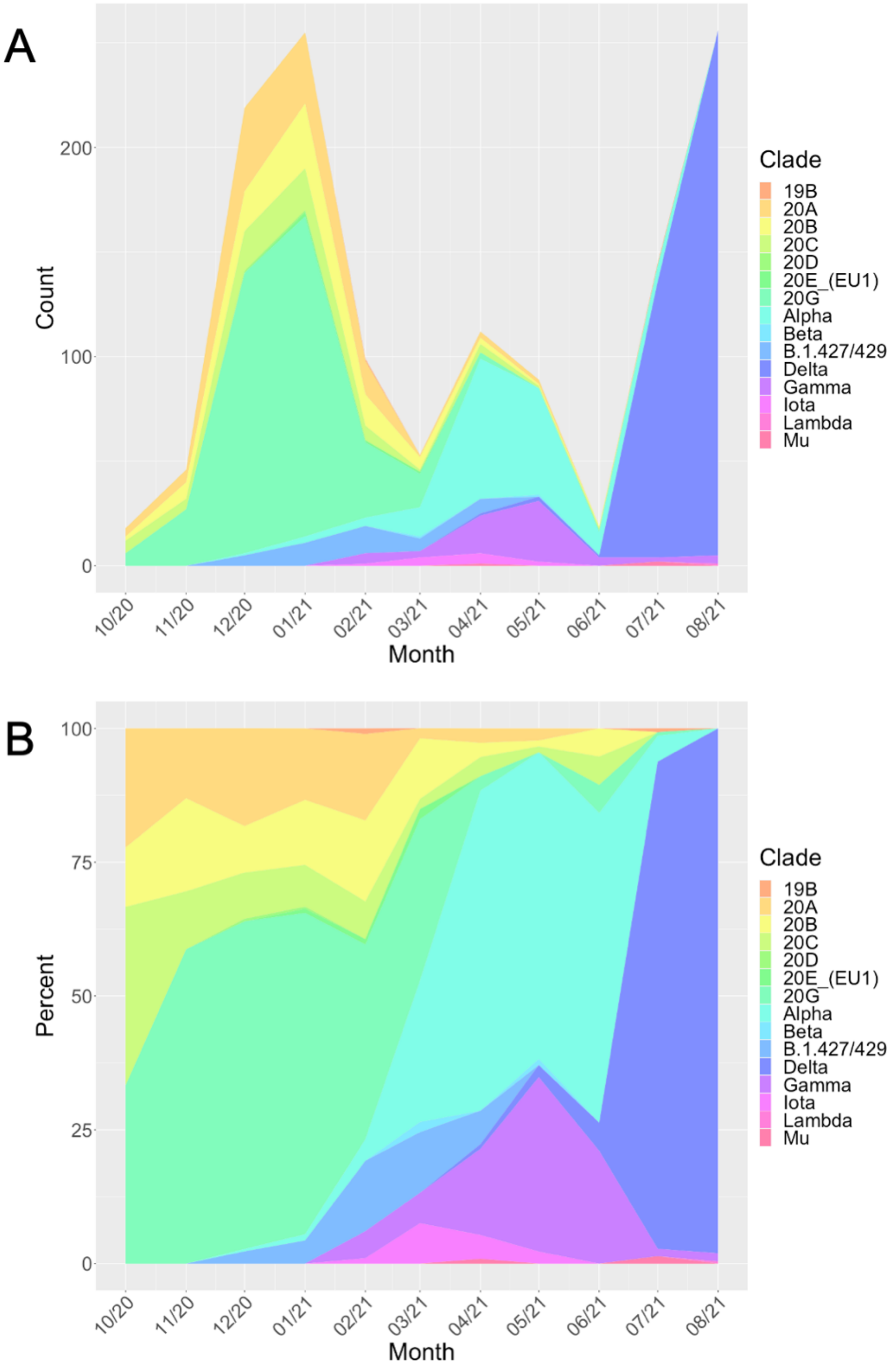

**Supplemental Tables**

**eTable 1: GISAID Accession Numbers, Lineages, and Variant of Concern (VOC) Classification of 499 SARS-CoV-2 Sequences Included in Study**

| ID | GISAID Accession | Pango Lineage | NextClade Clade | Variant Classification |
|---|---|---|---|---|
| LC_200 | EPI_ISL_1501823 | B.1.349 | 20C | Non-VOC |
| LC_201 | EPI_ISL_1501824 | B.1.582 | 20C | Non-VOC |
| LC_202 | EPI_ISL_1501825 | B.1.2 | 20G | Non-VOC |
| LC_203 | EPI_ISL_1501826 | B.1.2 | 20G | Non-VOC |
| LC_204 | EPI_ISL_1501827 | B.1.2 | 20G | Non-VOC |
| LC_205 | EPI_ISL_1501828 | B.1.2 | 20G | Non-VOC |
| LC_206 | EPI_ISL_1501829 | B.1.509 | 20C | Non-VOC |
| LC_207 | EPI_ISL_1501830 | B.1.2 | 20G | Non-VOC |
| LC_208 | EPI_ISL_1501831 | B.1.576 | 20C | Non-VOC |
| LC_209 | EPI_ISL_1501832 | B.1.2 | 20G | Non-VOC |
| LC_210 | EPI_ISL_1501833 | B.1.2 | 20G | Non-VOC |
| LC_211 | EPI_ISL_1501834 | B.1.2 | 20G | Non-VOC |
| LC_212 | EPI_ISL_1501835 | B.1.240 | 20A | Non-VOC |
| LC_213 | EPI_ISL_1501836 | B.1.243 | 20A | Non-VOC |
| LC_214 | EPI_ISL_1501837 | B.1.509 | 20C | Non-VOC |
| LC_215 | EPI_ISL_1501838 | B.1.2 | 20G | Non-VOC |
| LC_216 | EPI_ISL_1501839 | B.1.2 | 20G | Non-VOC |
| LC_217 | EPI_ISL_1501840 | B.1.240 | 20A | Non-VOC |
| LC_218 | EPI_ISL_1501841 | B.1.1.222 | 20B | Non-VOC |
| LC_219 | EPI_ISL_1501842 | B.1.2 | 20G | Non-VOC |
| LC_220 | EPI_ISL_1501843 | B.1.1 | 20B | Non-VOC |
| LC_221 | EPI_ISL_1501844 | B.1.1.518 | 20B | Non-VOC |
| LC_222 | EPI_ISL_1501845 | B.1.36.10 | 20A | Non-VOC |
| LC_223 | EPI_ISL_1501846 | B.1.139 | 20A | Non-VOC |
| LC_224 | EPI_ISL_1501847 | B.1.240 | 20A | Non-VOC |
| LC_225 | EPI_ISL_1501848 | B.1.2 | 20G | Non-VOC |
| LC_226 | EPI_ISL_1501849 | B.1.110.3 | 20A | Non-VOC |
| LC_227 | EPI_ISL_1501850 | B.1.2 | 20G | Non-VOC |
| LC_228 | EPI_ISL_1501851 | B.1.2 | 20G | Non-VOC |
| LC_229 | EPI_ISL_1501852 | B.1.2 | 20G | Non-VOC |
| LC_230 | EPI_ISL_1501853 | B.1.2 | 20G | Non-VOC |
| LC_231 | EPI_ISL_1501854 | B.1.2 | 20G | Non-VOC |
| LC_232 | EPI_ISL_1501855 | B.1.565 | 20A | Non-VOC |

| | | | | |
|---|---|---|---|---|
| LC_233 | EPI_ISL_1501856 | B.1.1.434 | 20B | Non-VOC |
| LC_234 | EPI_ISL_1501857 | B.1.2 | 20G | Non-VOC |
| LC_236 | EPI_ISL_1501858 | B.1.1.518 | 20B | Non-VOC |
| LC_237 | EPI_ISL_1501859 | B.1.2 | 20G | Non-VOC |
| LC_238 | EPI_ISL_1501860 | B.1.2 | 20G | Non-VOC |
| LC_239 | EPI_ISL_1501861 | B.1.2 | 20G | Non-VOC |
| LC_240 | EPI_ISL_1501862 | B.1 | 20A | Non-VOC |
| LC_241 | EPI_ISL_1501863 | B.1.2 | 20G | Non-VOC |
| LC_242 | EPI_ISL_1501864 | B.1.2 | 20G | Non-VOC |
| LC_243 | EPI_ISL_1501865 | B.1.2 | 20G | Non-VOC |
| LC_246 | EPI_ISL_1501866 | B.1.2 | 20G | Non-VOC |
| LC_248 | EPI_ISL_1501867 | B.1.2 | 20G | Non-VOC |
| LC_249 | EPI_ISL_1501868 | B.1.2 | 20G | Non-VOC |
| LC_250 | EPI_ISL_1501869 | B.1.2 | 20G | Non-VOC |
| LC_251 | EPI_ISL_1501870 | B.1.2 | 20G | Non-VOC |
| LC_252 | EPI_ISL_1501871 | C.31 | 20D | Non-VOC |
| LC_253 | EPI_ISL_1501872 | B.1.2 | 20G | Non-VOC |
| LC_254 | EPI_ISL_1501873 | B.1.2 | 20G | Non-VOC |
| LC_255 | EPI_ISL_1501874 | B.1.324 | 20C | Non-VOC |
| LC_256 | EPI_ISL_1501875 | B.1.2 | 20G | Non-VOC |
| LC_257 | EPI_ISL_1501876 | B.1.2 | 20G | Non-VOC |
| LC_258 | EPI_ISL_1501877 | B.1.2 | 20G | Non-VOC |
| LC_259 | EPI_ISL_1501878 | B.1.2 | 20G | Non-VOC |
| LC_260 | EPI_ISL_1501879 | B.1.2 | 20G | Non-VOC |
| LC_261 | EPI_ISL_1501880 | B.1.2 | 20G | Non-VOC |
| LC_262 | EPI_ISL_1501881 | B.1 | 20A | Non-VOC |
| LC_263 | EPI_ISL_1501882 | B.1.2 | 20G | Non-VOC |
| LC_264 | EPI_ISL_1501883 | B.1.2 | 20G | Non-VOC |
| LC_265 | EPI_ISL_1501884 | B.1.1 | 20B | Non-VOC |
| LC_266 | EPI_ISL_1501885 | B.1.2 | 20G | Non-VOC |
| LC_267 | EPI_ISL_1501886 | B.1.2 | 20G | Non-VOC |
| LC_268 | EPI_ISL_1501887 | B.1.2 | 20G | Non-VOC |
| LC_269 | EPI_ISL_1501888 | B.1.2 | 20G | Non-VOC |
| LC_270 | EPI_ISL_1501889 | B.1.565 | 20A | Non-VOC |
| LC_271 | EPI_ISL_1501890 | B.1.2 | 20G | Non-VOC |
| LC_272 | EPI_ISL_1501891 | B.1.2 | 20G | Non-VOC |
| LC_273 | EPI_ISL_1501892 | B.1.2 | 20G | Non-VOC |
| LC_274 | EPI_ISL_1501893 | B.1.234 | 20A | Non-VOC |
| LC_275 | EPI_ISL_1501894 | B.1.2 | 20G | Non-VOC |

| | | | | |
|---|---|---|---|---|
| LC_276 | EPI_ISL_1501895 | B.1.543 | 20A | Non-VOC |
| LC_277 | EPI_ISL_1501896 | B.1.2 | 20G | Non-VOC |
| LC_278 | EPI_ISL_1501897 | B.1.2 | 20G | Non-VOC |
| LC_279 | EPI_ISL_1501898 | B.1.1.222 | 20B | Non-VOC |
| LC_280 | EPI_ISL_1501899 | B.1.2 | 20G | Non-VOC |
| LC_281 | EPI_ISL_1501900 | B.1.2 | 20G | Non-VOC |
| LC_282 | EPI_ISL_1501901 | B.1.565 | 20A | Non-VOC |
| LC_283 | EPI_ISL_1501902 | B.1.2 | 20G | Non-VOC |
| LC_284 | EPI_ISL_1501903 | B.1.2 | 20G | Non-VOC |
| LC_285 | EPI_ISL_1501904 | B.1.1.222 | 20B | Non-VOC |
| LC_286 | EPI_ISL_1501905 | B.1.2 | 20G | Non-VOC |
| LC_287 | EPI_ISL_1501906 | B.1.2 | 20G | Non-VOC |
| LC_288 | EPI_ISL_1501907 | B.1.1.432 | 20B | Non-VOC |
| LC_289 | EPI_ISL_1501908 | B.1.2 | 20G | Non-VOC |
| LC_290 | EPI_ISL_1501909 | B.1.1.222 | 20B | Non-VOC |
| LC_291 | EPI_ISL_1501910 | B.1.2 | 20G | Non-VOC |
| LC_292 | EPI_ISL_1501911 | B.1.243 | 20A | Non-VOC |
| LC_293 | EPI_ISL_1501912 | B.1.596 | 20G | Non-VOC |
| LC_294 | EPI_ISL_1501913 | B.1.2 | 20G | Non-VOC |
| LC_295 | EPI_ISL_1501914 | B.1.2 | 20G | Non-VOC |
| LC_296 | EPI_ISL_1501915 | B.1.2 | 20G | Non-VOC |
| LC_297 | EPI_ISL_1501916 | B.1.1 | 20B | Non-VOC |
| LC_299 | EPI_ISL_1501918 | B.1 | 20A | Non-VOC |
| LC_300 | EPI_ISL_1501919 | B.1.1.434 | 20B | Non-VOC |
| LC_301 | EPI_ISL_1501920 | B.1.576 | 20C | Non-VOC |
| LC_303 | EPI_ISL_1501921 | B.1.139 | 20A | Non-VOC |
| LC_305 | EPI_ISL_1501922 | B.1.2 | 20G | Non-VOC |
| LC_306 | EPI_ISL_1501923 | B.1.2 | 20G | Non-VOC |
| LC_307 | EPI_ISL_1501924 | B.1.2 | 20G | Non-VOC |
| LC_308 | EPI_ISL_1501925 | B.1.2 | 20G | Non-VOC |
| LC_310 | EPI_ISL_1501927 | B.1.305 | 20C | Non-VOC |
| LC_311 | EPI_ISL_1501928 | B.1.1.518 | 20B | Non-VOC |
| LC_312 | EPI_ISL_1501929 | B.1.396 | 20A | Non-VOC |
| LC_314 | EPI_ISL_1501930 | C.31 | 20D | Non-VOC |
| LC_315 | EPI_ISL_1501931 | B.1.2 | 20G | Non-VOC |
| LC_316 | EPI_ISL_1501932 | B.1.576 | 20C | Non-VOC |
| LC_317 | EPI_ISL_1501933 | B.1 | 20B | Non-VOC |
| LC_318 | EPI_ISL_1501934 | B.1.2 | 20G | Non-VOC |
| LC_319 | EPI_ISL_1501935 | B.1.2 | 20G | Non-VOC |

| | | | | |
|---|---|---|---|---|
| LC_320 | EPI_ISL_1501936 | B.1.2 | 20G | Non-VOC |
| LC_321 | EPI_ISL_1501937 | B.1.2 | 20G | Non-VOC |
| LC_322 | EPI_ISL_1501938 | B.1.2 | 20G | Non-VOC |
| LC_323 | EPI_ISL_1501939 | B.1.1.518 | 20B | Non-VOC |
| LC_324 | EPI_ISL_1501940 | B.1.1.416 | 20B | Non-VOC |
| LC_325 | EPI_ISL_1501941 | B.1.595 | 20C | Non-VOC |
| LC_326 | EPI_ISL_1501942 | B.1.2 | 20G | Non-VOC |
| LC_327 | EPI_ISL_1501943 | B.1.578 | 20C | Non-VOC |
| LC_328 | EPI_ISL_1501944 | B.1 | 20A | Non-VOC |
| LC_329 | EPI_ISL_1501945 | B.1.2 | 20G | Non-VOC |
| LC_330 | EPI_ISL_1501946 | B.1.2 | 20G | Non-VOC |
| LC_331 | EPI_ISL_1501947 | B.1.2 | 20G | Non-VOC |
| LC_332 | EPI_ISL_1501948 | B.1.2 | 20G | Non-VOC |
| LC_333 | EPI_ISL_1501949 | B.1.2 | 20G | Non-VOC |
| LC_334 | EPI_ISL_1501950 | B.1.565 | 20A | Non-VOC |
| LC_335 | EPI_ISL_1501951 | B.1.509 | 20C | Non-VOC |
| LC_336 | EPI_ISL_1501952 | B.1.2 | 20G | Non-VOC |
| LC_338 | EPI_ISL_1501953 | B.1.1.317 | 20B | Non-VOC |
| LC_339 | EPI_ISL_1501954 | B.1.2 | 20G | Non-VOC |
| LC_340 | EPI_ISL_1501955 | B.1.2 | 20G | Non-VOC |
| LC_341 | EPI_ISL_1501956 | B.1.427 | 20C | Non-VOC |
| LC_342 | EPI_ISL_1501957 | B.1.232 | 20A | Non-VOC |
| LC_346 | EPI_ISL_1501959 | B.1.2 | 20G | Non-VOC |
| LC_347 | EPI_ISL_1501960 | B.1.1.192 | 20B | Non-VOC |
| LC_348 | EPI_ISL_1501961 | B.1.2 | 20G | Non-VOC |
| LC_349 | EPI_ISL_1501962 | B.1.2 | 20G | Non-VOC |
| LC_350 | EPI_ISL_1501963 | B.1.240 | 20A | Non-VOC |
| LC_351 | EPI_ISL_1501964 | A.2.5 | 19B | Non-VOC |
| LC_352 | EPI_ISL_1501965 | B.1.2 | 20G | Non-VOC |
| LC_353 | EPI_ISL_1501966 | B.1.2 | 20G | Non-VOC |
| LC_354 | EPI_ISL_1501967 | B.1.232 | 20A | Non-VOC |
| LC_355 | EPI_ISL_1501968 | B.1.139 | 20A | Non-VOC |
| LC_356 | EPI_ISL_1501969 | B.1.576 | 20C | Non-VOC |
| LC_357 | EPI_ISL_1501970 | B.1.2 | 20G | Non-VOC |
| LC_359 | EPI_ISL_1501972 | B.1 | 20C | Non-VOC |
| LC_360 | EPI_ISL_1501973 | B.1.2 | 20G | Non-VOC |
| LC_361 | EPI_ISL_1501974 | B.1.2 | 20G | Non-VOC |
| LC_362 | EPI_ISL_1501975 | B.1.429 | 20C | Non-VOC |
| LC_363 | EPI_ISL_1501976 | B.1.2 | 20G | Non-VOC |

| | | | | |
|---|---|---|---|---|
| LC_365 | EPI_ISL_1501977 | B.1.2 | 20G | Non-VOC |
| LC_367 | EPI_ISL_1501978 | B.1.2 | 20G | Non-VOC |
| LC_368 | EPI_ISL_1501979 | B.1.2 | 20G | Non-VOC |
| LC_370 | EPI_ISL_1501980 | B.1.2 | 20G | Non-VOC |
| LC_371 | EPI_ISL_1501981 | B.1.427 | 20C | Non-VOC |
| LC_373 | EPI_ISL_1501983 | B.1.1.7 | 20I/501Y.V1 | Alpha |
| LC_374 | EPI_ISL_1501984 | B.1.2 | 20G | Non-VOC |
| LC_375 | EPI_ISL_1501985 | B.1.2 | 20G | Non-VOC |
| LC_376 | EPI_ISL_1501986 | B.1.2 | 20G | Non-VOC |
| LC_377 | EPI_ISL_1501987 | B.1.2 | 20G | Non-VOC |
| LC_378 | EPI_ISL_1501988 | B.1.1.7 | 20I/501Y.V1 | Alpha |
| LC_379 | EPI_ISL_1501989 | B.1.1.518 | 20B | Non-VOC |
| LC_380 | EPI_ISL_1501990 | B.1.2 | 20G | Non-VOC |
| LC_381 | EPI_ISL_1501991 | B.1.232 | 20A | Non-VOC |
| LC_382 | EPI_ISL_1501992 | B.1.596 | 20G | Non-VOC |
| LC_383 | EPI_ISL_1501993 | B.1 | 20C | Non-VOC |
| LC_384 | EPI_ISL_1501994 | B.1.429 | 20C | Non-VOC |
| LC_385 | EPI_ISL_1501995 | B.1.239 | 20A | Non-VOC |
| LC_386 | EPI_ISL_1501996 | B.1.1.519 | 20B | Non-VOC |
| LC_387 | EPI_ISL_1501997 | B.1.427 | 20C | Non-VOC |
| LC_388 | EPI_ISL_1501998 | B.1.1.222 | 20B | Non-VOC |
| LC_389 | EPI_ISL_1501999 | B.1.2 | 20G | Non-VOC |
| LC_390 | EPI_ISL_1502000 | B.1.243 | 20A | Non-VOC |
| LC_391 | EPI_ISL_1502001 | B.1.2 | 20G | Non-VOC |
| LC_392 | EPI_ISL_1502002 | B.1.2 | 20G | Non-VOC |
| LC_393 | EPI_ISL_1502003 | B.1.2 | 20G | Non-VOC |
| LC_394 | EPI_ISL_1502004 | B.1.609 | 20A | Non-VOC |
| LC_396 | EPI_ISL_1502005 | B.1.427 | 20C | Non-VOC |
| LC_397 | EPI_ISL_1502006 | B.1.427 | 20C | Non-VOC |
| LC_398 | EPI_ISL_1502007 | B.1.429 | 20C | Non-VOC |
| LC_399 | EPI_ISL_1502008 | B.1.1.7 | 20I/501Y.V1 | Alpha |
| LC_400 | EPI_ISL_1502009 | B.1.2 | 20G | Non-VOC |
| LC_401 | EPI_ISL_1502010 | B.1.2 | 20G | Non-VOC |
| LC_403 | EPI_ISL_1502011 | B.1.427 | 20C | Non-VOC |
| LC_404 | EPI_ISL_1502012 | B.1.2 | 20G | Non-VOC |
| LC_405 | EPI_ISL_1502013 | B.1.1.7 | 20I/501Y.V1 | Alpha |
| LC_406 | EPI_ISL_1502014 | B.1.2 | 20G | Non-VOC |
| LC_409 | EPI_ISL_1502015 | B.1.2 | 20G | Non-VOC |
| LC_410 | EPI_ISL_1502016 | B.1.2 | 20G | Non-VOC |

| | | | | |
|---|---|---|---|---|
| LC_411 | EPI_ISL_1502017 | B.1.2 | 20G | Non-VOC |
| LC_412 | EPI_ISL_1502018 | B.1.243 | 20A | Non-VOC |
| LC_413 | EPI_ISL_1502019 | B.1.2 | 20G | Non-VOC |
| LC_414 | EPI_ISL_1502020 | B.1.400 | 20A | Non-VOC |
| LC_415 | EPI_ISL_1502021 | P.1 | 20J/501Y.V3 | Gamma |
| LC_416 | EPI_ISL_1502022 | B.1.400 | 20A | Non-VOC |
| LC_417 | EPI_ISL_1502023 | B.1.243 | 20A | Non-VOC |
| LC_418 | EPI_ISL_1502024 | B.1.2 | 20G | Non-VOC |
| LC_420 | EPI_ISL_1502025 | P.2 | 20B | Non-VOC |
| LC_421 | EPI_ISL_1502026 | B.1.1.222 | 20B | Non-VOC |
| LC_422 | EPI_ISL_1502027 | P.1 | 20J/501Y.V3 | Gamma |
| LC_423 | EPI_ISL_1502028 | B.1.239 | 20A | Non-VOC |
| LC_424 | EPI_ISL_1502029 | B.1.1.519 | 20B | Non-VOC |
| LC_425 | EPI_ISL_1502030 | B.1.1.7 | 20I/501Y.V1 | Alpha |
| LC_426 | EPI_ISL_1502031 | B.1.2 | 20G | Non-VOC |
| LC_427 | EPI_ISL_1502032 | B.1.1.7 | 20I/501Y.V1 | Alpha |
| LC_428 | EPI_ISL_1502033 | B.1.2 | 20G | Non-VOC |
| LC_429 | EPI_ISL_1502034 | P.2 | 20B | Non-VOC |
| LC_430 | EPI_ISL_1502035 | B.1.2 | 20G | Non-VOC |
| LC_431 | EPI_ISL_1502036 | B.1.1.7 | 20I/501Y.V1 | Alpha |
| LC_432 | EPI_ISL_1502037 | B.1.427 | 20C | Non-VOC |
| LC_433 | EPI_ISL_1502038 | B.1.1.7 | 20I/501Y.V1 | Alpha |
| LC_434 | EPI_ISL_1502039 | B.1.2 | 20G | Non-VOC |
| LC_435 | EPI_ISL_1502040 | P.1 | 20J/501Y.V3 | Gamma |
| LC_436 | EPI_ISL_1502041 | B.1.1.7 | 20I/501Y.V1 | Alpha |
| LC_437 | EPI_ISL_1502042 | B.1.1.7 | 20I/501Y.V1 | Alpha |
| LC_438 | EPI_ISL_1502043 | B.1.427 | 20C | Non-VOC |
| LC_439 | EPI_ISL_1502044 | B.1.427 | 20C | Non-VOC |
| LC_440 | EPI_ISL_1502045 | B.1.1.519 | 20B | Non-VOC |
| LC_441 | EPI_ISL_1502046 | B.1.427 | 20C | Non-VOC |
| LC_442 | EPI_ISL_1502047 | B.1.427 | 20C | Non-VOC |
| LC_443 | EPI_ISL_1704763 | B.1.1.7 | 20I/501Y.V1 | Alpha |
| LC_444 | EPI_ISL_1704764 | B.1.1.7 | 20I/501Y.V1 | Alpha |
| LC_445 | EPI_ISL_1704765 | B.1.429 | 20C | Non-VOC |
| LC_446 | EPI_ISL_1704766 | B.1.429 | 20C | Non-VOC |
| LC_447 | EPI_ISL_1704767 | B.1.2 | 20G | Non-VOC |
| LC_448 | EPI_ISL_1704768 | B.1.2 | 20G | Non-VOC |
| LC_449 | EPI_ISL_1704769 | B.1.2 | 20G | Non-VOC |
| LC_450 | EPI_ISL_1704770 | B.1.526 | 20C | Non-VOC |

| | | | | |
|---|---|---|---|---|
| LC_451 | EPI_ISL_1704771 | B.1.1.7 | 20I/501Y.V1 | Alpha |
| LC_452 | EPI_ISL_1704772 | P.1 | 20J/501Y.V3 | Gamma |
| LC_453 | EPI_ISL_1704773 | B.1.1.7 | 20I/501Y.V1 | Alpha |
| LC_454 | EPI_ISL_1704715 | B.1.2 | 20G | Non-VOC |
| LC_455 | EPI_ISL_1704716 | B.1.429 | 20C | Non-VOC |
| LC_456 | EPI_ISL_1704717 | B.1.427 | 20C | Non-VOC |
| LC_457 | EPI_ISL_1704718 | B.1.427 | 20C | Non-VOC |
| LC_458 | EPI_ISL_1704719 | B.1.427 | 20C | Non-VOC |
| LC_459 | EPI_ISL_1704720 | B.1.1.519 | 20B | Non-VOC |
| LC_460 | EPI_ISL_1704721 | B.1.1.519 | 20B | Non-VOC |
| LC_461 | EPI_ISL_1704722 | R.1 | 20B | Non-VOC |
| LC_462 | EPI_ISL_1704723 | B.1.1.7 | 20I/501Y.V1 | Alpha |
| LC_463 | EPI_ISL_1704724 | B.1.2 | 20G | Non-VOC |
| LC_464 | EPI_ISL_1704725 | B.1.2 | 20G | Non-VOC |
| LC_465 | EPI_ISL_1704726 | B.1.1.7 | 20I/501Y.V1 | Alpha |
| LC_466 | EPI_ISL_1704727 | B.1.429 | 20C | Non-VOC |
| LC_467 | EPI_ISL_1704728 | B.1.1.7 | 20I/501Y.V1 | Alpha |
| LC_468 | EPI_ISL_1704729 | P.1 | 20J/501Y.V3 | Gamma |
| LC_469 | EPI_ISL_1704730 | B.1.1.7 | 20I/501Y.V1 | Alpha |
| LC_470 | EPI_ISL_2009368 | B.1.2 | 20G | Non-VOC |
| LC_471 | EPI_ISL_2009369 | P.1 | 20J/501Y.V3 | Gamma |
| LC_472 | EPI_ISL_2009370 | Q.3 | 20I/501Y.V1 | Alpha |
| LC_473 | EPI_ISL_2009371 | P.1 | 20J/501Y.V3 | Gamma |
| LC_474 | EPI_ISL_2009372 | B.1.1.7 | 20I/501Y.V1 | Alpha |
| LC_475 | EPI_ISL_2009373 | B.1.1.7 | 20I/501Y.V1 | Alpha |
| LC_476 | EPI_ISL_2009374 | B.1.1.7 | 20I/501Y.V1 | Alpha |
| LC_477 | EPI_ISL_2009375 | B.1.1.7 | 20I/501Y.V1 | Alpha |
| LC_478 | EPI_ISL_1704731 | B.1.429 | 20C | Non-VOC |
| LC_479 | EPI_ISL_1704732 | B.1.351 | 20H/501Y.V2 | Beta |
| LC_480 | EPI_ISL_1704733 | B.1.351 | 20H/501Y.V2 | Beta |
| LC_481 | EPI_ISL_1704734 | B.1.1.7 | 20I/501Y.V1 | Alpha |
| LC_482 | EPI_ISL_1704735 | B.1.596 | 20G | Non-VOC |
| LC_483 | EPI_ISL_1704736 | B.1.1.7 | 20I/501Y.V1 | Alpha |
| LC_484 | EPI_ISL_1704737 | B.1.1.7 | 20I/501Y.V1 | Alpha |
| LC_485 | EPI_ISL_1704738 | P.1 | 20J/501Y.V3 | Gamma |
| LC_486 | EPI_ISL_1704739 | B.1.1.7 | 20I/501Y.V1 | Alpha |
| LC_487 | EPI_ISL_1704740 | B.1.526 | 20C | Non-VOC |
| LC_488 | EPI_ISL_1704741 | P.1 | 20J/501Y.V3 | Gamma |
| LC_489 | EPI_ISL_1704742 | B.1.1.7 | 20I/501Y.V1 | Alpha |

| | | | | |
|---|---|---|---|---|
| LC_490 | EPI_ISL_1704743 | B.1.1.7 | 20I/501Y.V1 | Alpha |
| LC_491 | EPI_ISL_1704744 | P.1 | 20J/501Y.V3 | Gamma |
| LC_492 | EPI_ISL_1704745 | B.1.1.7 | 20I/501Y.V1 | Alpha |
| LC_493 | EPI_ISL_1704746 | B.1.1.7 | 20I/501Y.V1 | Alpha |
| LC_494 | EPI_ISL_1704747 | B.1.1.7 | 20I/501Y.V1 | Alpha |
| LC_495 | EPI_ISL_1704748 | B.1.1.7 | 20I/501Y.V1 | Alpha |
| LC_496 | EPI_ISL_1704749 | B.1.1.7 | 20I/501Y.V1 | Alpha |
| LC_497 | EPI_ISL_1704750 | B.1.2 | 20G | Non-VOC |
| LC_498 | EPI_ISL_1704751 | P.1 | 20J/501Y.V3 | Gamma |
| LC_499 | EPI_ISL_1704752 | B.1.427 | 20C | Non-VOC |
| LC_500 | EPI_ISL_1704753 | B.1.427 | 20C | Non-VOC |
| LC_501 | EPI_ISL_1704754 | B.1.1.7 | 20I/501Y.V1 | Alpha |
| LC_502 | EPI_ISL_1704755 | B.1.1.7 | 20I/501Y.V1 | Alpha |
| LC_503 | EPI_ISL_1704756 | P.1.12 | 20J/501Y.V3 | Gamma |
| LC_504 | EPI_ISL_1704757 | B.1.1.7 | 20I/501Y.V1 | Alpha |
| LC_505 | EPI_ISL_1704758 | B.1.427 | 20C | Non-VOC |
| LC_506 | EPI_ISL_1704759 | B.1.429 | 20C | Non-VOC |
| LC_507 | EPI_ISL_1704760 | R.1 | 20B | Non-VOC |
| LC_508 | EPI_ISL_1704761 | R.1 | 20B | Non-VOC |
| LC_509 | EPI_ISL_1704762 | B.1.1.7 | 20I/501Y.V1 | Alpha |
| LC_510 | EPI_ISL_2009376 | B.1.1.7 | 20I/501Y.V1 | Alpha |
| LC_512 | EPI_ISL_2009377 | B.1.1.7 | 20I/501Y.V1 | Alpha |
| LC_513 | EPI_ISL_2009378 | P.2 | 20B | Non-VOC |
| LC_515 | EPI_ISL_2009380 | B.1.1.7 | 20I/501Y.V1 | Alpha |
| LC_516 | EPI_ISL_2009381 | B.1.637 | 20C | Non-VOC |
| LC_517 | EPI_ISL_2009382 | B.1.1.7 | 20I/501Y.V1 | Alpha |
| LC_519 | EPI_ISL_2009383 | B.1.1.7 | 20I/501Y.V1 | Alpha |
| LC_520 | EPI_ISL_2009384 | B.1.1.7 | 20I/501Y.V1 | Alpha |
| LC_521 | EPI_ISL_2009385 | B.1.1.7 | 20I/501Y.V1 | Alpha |
| LC_523 | EPI_ISL_2009386 | B.1.429 | 20C | Non-VOC |
| LC_524 | EPI_ISL_2009387 | B.1.1.7 | 20I/501Y.V1 | Alpha |
| LC_525 | EPI_ISL_2009388 | B.1.1.7 | 20I/501Y.V1 | Alpha |
| LC_526 | EPI_ISL_2009389 | P.1 | 20J/501Y.V3 | Gamma |
| LC_527 | EPI_ISL_2009390 | B.1.1.7 | 20I/501Y.V1 | Alpha |
| LC_528 | EPI_ISL_2009391 | B.1.1.7 | 20I/501Y.V1 | Alpha |
| LC_529 | EPI_ISL_2009392 | B.1.351 | 20H/501Y.V2 | Beta |
| LC_531 | EPI_ISL_2009394 | P.1 | 20J/501Y.V3 | Gamma |
| LC_532 | EPI_ISL_2009395 | P.1 | 20J/501Y.V3 | Gamma |
| LC_578 | EPI_ISL_2373051 | B.1.1.7 | 20I/501Y.V1 | Alpha |

| | | | | |
|---|---|---|---|---|
| LC_580 | EPI_ISL_2373052 | B.1.1.519 | 20B | Non-VOC |
| LC_581 | EPI_ISL_2373053 | B.1.1.7 | 20I/501Y.V1 | Alpha |
| LC_582 | EPI_ISL_2373054 | B.1.1.7 | 20I/501Y.V1 | Alpha |
| LC_583 | EPI_ISL_2373055 | P.1 | 20J/501Y.V3 | Gamma |
| LC_584 | EPI_ISL_2373056 | B.1.1.7 | 20I/501Y.V1 | Alpha |
| LC_586 | EPI_ISL_2373057 | P.1 | 20J/501Y.V3 | Gamma |
| LC_587 | EPI_ISL_2373058 | Q.3 | 20I/501Y.V1 | Alpha |
| LC_590 | EPI_ISL_2373059 | B.1.1.7 | 20I/501Y.V1 | Alpha |
| LC_591 | EPI_ISL_2373060 | B.1.1.7 | 20I/501Y.V1 | Alpha |
| LC_592 | EPI_ISL_2373061 | P.1 | 20J/501Y.V3 | Gamma |
| LC_593 | EPI_ISL_2373062 | P.1 | 20J/501Y.V3 | Gamma |
| LC_594 | EPI_ISL_2373063 | B.1.637 | 20C | Non-VOC |
| LC_595 | EPI_ISL_2373064 | P.1 | 20J/501Y.V3 | Gamma |
| LC_596 | EPI_ISL_2373065 | B.1.1.7 | 20I/501Y.V1 | Alpha |
| LC_597 | EPI_ISL_2373066 | B.1.1.7 | 20I/501Y.V1 | Alpha |
| LC_598 | EPI_ISL_2373067 | B.1.1.7 | 20I/501Y.V1 | Alpha |
| LC_599 | EPI_ISL_2373068 | B.1.1.519 | 20B | Non-VOC |
| LC_600 | EPI_ISL_2373069 | P.1 | 20J/501Y.V3 | Gamma |
| LC_601 | EPI_ISL_2373070 | P.1 | 20J/501Y.V3 | Gamma |
| LC_604 | EPI_ISL_2373071 | None | 20I/501Y.V1 | Alpha |
| LC_605 | EPI_ISL_2373072 | B.1.1.7 | 20I/501Y.V1 | Alpha |
| LC_606 | EPI_ISL_2373073 | B.1.1.7 | 20I/501Y.V1 | Alpha |
| LC_607 | EPI_ISL_2373074 | B.1.1.7 | 20I/501Y.V1 | Alpha |
| LC_608 | EPI_ISL_2373075 | B.1.1.7 | 20I/501Y.V1 | Alpha |
| LC_609 | EPI_ISL_2373076 | P.1 | 20J/501Y.V3 | Gamma |
| LC_610 | EPI_ISL_2373077 | B.1.1.222 | 20B | Non-VOC |
| LC_611 | EPI_ISL_2373078 | B.1.1.7 | 20I/501Y.V1 | Alpha |
| LC_612 | EPI_ISL_2373079 | P.1 | 20J/501Y.V3 | Gamma |
| LC_613 | EPI_ISL_2373080 | B.1.1.7 | 20I/501Y.V1 | Alpha |
| LC_614 | EPI_ISL_2373081 | P.1 | 20J/501Y.V3 | Gamma |
| LC_615 | EPI_ISL_2373082 | B.1.1.7 | 20I/501Y.V1 | Alpha |
| LC_616 | EPI_ISL_2373083 | B.1.1.7 | 20I/501Y.V1 | Alpha |
| LC_617 | EPI_ISL_2373084 | B.1.1.7 | 20I/501Y.V1 | Alpha |
| LC_618 | EPI_ISL_2373085 | B.1.1.7 | 20I/501Y.V1 | Alpha |
| LC_619 | EPI_ISL_2373086 | B.1.525 | 20A | Non-VOC |
| LC_620 | EPI_ISL_2373087 | B.1.1.7 | 20I/501Y.V1 | Alpha |
| LC_621 | EPI_ISL_2373088 | B.1.1.7 | 20I/501Y.V1 | Alpha |
| LC_622 | EPI_ISL_2373089 | B.1.1.7 | 20I/501Y.V1 | Alpha |
| LC_623 | EPI_ISL_2373090 | B.1.1.7 | 20I/501Y.V1 | Alpha |

| | | | | |
|---|---|---|---|---|
| LC_624 | EPI_ISL_2373091 | B.1.1.7 | 20I/501Y.V1 | Alpha |
| LC_625 | EPI_ISL_2373092 | B.1.1.7 | 20I/501Y.V1 | Alpha |
| LC_626 | EPI_ISL_2373093 | B.1.1.7 | 20I/501Y.V1 | Alpha |
| LC_627 | EPI_ISL_2373094 | P.1 | 20J/501Y.V3 | Gamma |
| LC_628 | EPI_ISL_2373095 | B.1.1.7 | 20I/501Y.V1 | Alpha |
| LC_629 | EPI_ISL_2373096 | B.1.1.7 | 20I/501Y.V1 | Alpha |
| LC_630 | EPI_ISL_2373097 | B.1.1.7 | 20I/501Y.V1 | Alpha |
| LC_631 | EPI_ISL_2373098 | B.1.1.7 | 20I/501Y.V1 | Alpha |
| LC_632 | EPI_ISL_2373099 | B.1.1.7 | 20I/501Y.V1 | Alpha |
| LC_633 | EPI_ISL_2373100 | P.1 | 20J/501Y.V3 | Gamma |
| LC_634 | EPI_ISL_2373101 | B.1.1.7 | 20I/501Y.V1 | Alpha |
| LC_635 | EPI_ISL_2373102 | B.1.1.7 | 20I/501Y.V1 | Alpha |
| LC_636 | EPI_ISL_2373103 | P.1 | 20J/501Y.V3 | Gamma |
| LC_637 | EPI_ISL_2373104 | P.1 | 20J/501Y.V3 | Gamma |
| LC_638 | EPI_ISL_2373105 | B.1.1.7 | 20I/501Y.V1 | Alpha |
| LC_639 | EPI_ISL_2373106 | B.1.1.7 | 20I/501Y.V1 | Alpha |
| LC_640 | EPI_ISL_2373107 | P.1 | 20J/501Y.V3 | Gamma |
| LC_641 | EPI_ISL_2842585 | P.1 | 20J (Gamma, V3) | Gamma |
| LC_642 | EPI_ISL_2842586 | None | 20I (Alpha, V1) | Alpha |
| LC_643 | EPI_ISL_2842587 | P.1 | 20J (Gamma, V3) | Gamma |
| LC_644 | EPI_ISL_2842588 | B.1.1.7 | 20I (Alpha, V1) | Alpha |
| LC_645 | EPI_ISL_2842589 | P.1 | 20J (Gamma, V3) | Gamma |
| LC_646 | EPI_ISL_2842590 | B.1.1.7 | 20I (Alpha, V1) | Alpha |
| LC_647 | EPI_ISL_2842591 | B.1.1.7 | 20I (Alpha, V1) | Alpha |
| LC_648 | EPI_ISL_2842592 | B.1.1.7 | 20I (Alpha, V1) | Alpha |
| LC_649 | EPI_ISL_2842593 | B.1.1.7 | 20I (Alpha, V1) | Alpha |
| LC_650 | EPI_ISL_2842594 | P.1 | 20J (Gamma, V3) | Gamma |
| LC_651 | EPI_ISL_2842595 | B.1.1.7 | 20I (Alpha, V1) | Alpha |
| LC_654 | EPI_ISL_2842580 | AY.5 | 21A (Delta) | Delta |
| LC_655 | EPI_ISL_2842581 | P.1 | 20J (Gamma, V3) | Gamma |
| LC_657 | EPI_ISL_2842582 | P.1 | 20J (Gamma, V3) | Gamma |
| LC_658 | EPI_ISL_2842583 | P.1 | 20J (Gamma, V3) | Gamma |

| | | | | |
|---|---|---|---|---|
| LC_659 | EPI_ISL_2842584 | B.1.1.7 | 20I (Alpha, V1) | Alpha |
| LC_660 | EPI_ISL_3304311 | AY.4 | 21A (Delta) | Delta |
| LC_661 | EPI_ISL_3304312 | B.1.1.7 | 20I (Alpha, V1) | Alpha |
| LC_662 | EPI_ISL_3304313 | B.1.617.2 | 21A (Delta) | Delta |
| LC_663 | EPI_ISL_3185847 | P.1 | 20J (Gamma, V3) | Gamma |
| LC_664 | EPI_ISL_3185848 | AY.3 | 21A (Delta) | Delta |
| LC_665 | EPI_ISL_3185849 | AY.3 | 21A (Delta) | Delta |
| LC_666 | EPI_ISL_3185850 | AY.25 | 21A (Delta) | Delta |
| LC_667 | EPI_ISL_3185851 | AY.4 | 21A (Delta) | Delta |
| LC_671 | EPI_ISL_3304314 | AY.23 | 21A (Delta) | Delta |
| LC_672 | EPI_ISL_3304315 | AY.4 | 21A (Delta) | Delta |
| LC_673 | EPI_ISL_3304316 | B.1.617.2 | 21A (Delta) | Delta |
| LC_674 | EPI_ISL_3304317 | AY.5 | 21A (Delta) | Delta |
| LC_675 | EPI_ISL_3304318 | AY.25 | 21A (Delta) | Delta |
| LC_677 | EPI_ISL_3304319 | B.1.617.2 | 21A (Delta) | Delta |
| LC_679 | EPI_ISL_3304320 | AY.4 | 21A (Delta) | Delta |
| LC_680 | EPI_ISL_3304321 | AY.4 | 21A (Delta) | Delta |
| LC_681 | EPI_ISL_3304322 | AY.25 | 21A (Delta) | Delta |
| LC_682 | EPI_ISL_3304323 | AY.4 | 21A (Delta) | Delta |
| LC_683 | EPI_ISL_3304324 | B.1.617.2 | 21A (Delta) | Delta |
| LC_684 | EPI_ISL_3304325 | B.1.617.2 | 21A (Delta) | Delta |
| LC_685 | EPI_ISL_3304326 | AY.4 | 21A (Delta) | Delta |
| LC_686 | EPI_ISL_3304327 | B.1.1.7 | 20I (Alpha, V1) | Alpha |
| LC_687 | EPI_ISL_3304328 | AY.20 | 21A (Delta) | Delta |
| LC_688 | EPI_ISL_3304329 | B.1.617.2 | 21A (Delta) | Delta |
| LC_689 | EPI_ISL_3304330 | B.1.617.2 | 21A (Delta) | Delta |
| LC_690 | EPI_ISL_3304331 | AY.3 | 21A (Delta) | Delta |
| LC_691 | EPI_ISL_3304332 | AY.26 | 21A (Delta) | Delta |
| LC_692 | EPI_ISL_3304333 | B.1.617.2 | 21A (Delta) | Delta |
| LC_693 | EPI_ISL_3304334 | B.1.617.2 | 21A (Delta) | Delta |
| LC_694 | EPI_ISL_3304335 | AY.4 | 21A (Delta) | Delta |
| LC_695 | EPI_ISL_3304336 | AY.3 | 21A (Delta) | Delta |
| LC_697 | EPI_ISL_3304338 | B.1.617.2 | 21A (Delta) | Delta |
| LC_698 | EPI_ISL_3304339 | AY.10 | 21A (Delta) | Delta |
| LC_700 | EPI_ISL_3304340 | AY.4 | 21A (Delta) | Delta |
| LC_701 | EPI_ISL_3304341 | AY.3 | 21A (Delta) | Delta |
| LC_702 | EPI_ISL_3304342 | B.1.617.2 | 21A (Delta) | Delta |

| | | | | |
|---|---|---|---|---|
| LC_703 | EPI_ISL_3304343 | B.1.617.2 | 21A (Delta) | Delta |
| LC_704 | EPI_ISL_3304344 | B.1.617.2 | 21A (Delta) | Delta |
| LC_705 | EPI_ISL_4026596 | B.1.617.2 | 21A (Delta) | Delta |
| LC_706 | EPI_ISL_4026597 | AY.3 | 21A (Delta) | Delta |
| LC_707 | EPI_ISL_4026598 | AY.3 | 21A (Delta) | Delta |
| LC_708 | EPI_ISL_4026599 | B.1.617.2 | 21A (Delta) | Delta |
| LC_709 | EPI_ISL_4026600 | B.1.617.2 | 21A (Delta) | Delta |
| LC_710 | EPI_ISL_4026601 | AY.26 | 21A (Delta) | Delta |
| LC_711 | EPI_ISL_4026602 | AY.25 | 21A (Delta) | Delta |
| LC_712 | EPI_ISL_4026603 | AY.3 | 21A (Delta) | Delta |
| LC_713 | EPI_ISL_4026604 | B.1.617.2 | 21A (Delta) | Delta |
| LC_714 | EPI_ISL_4026605 | B.1.617.2 | 21A (Delta) | Delta |
| LC_715 | EPI_ISL_4026606 | AY.25 | 21A (Delta) | Delta |
| LC_716 | EPI_ISL_4026607 | B.1.617.2 | 21A (Delta) | Delta |
| LC_717 | EPI_ISL_4026608 | AY.3.1 | 21A (Delta) | Delta |
| LC_718 | EPI_ISL_4026609 | AY.3 | 21A (Delta) | Delta |
| LC_719 | EPI_ISL_4026610 | AY.25 | 21A (Delta) | Delta |
| LC_720 | EPI_ISL_4026611 | AY.25 | 21A (Delta) | Delta |
| LC_721 | EPI_ISL_4026612 | AY.3 | 21A (Delta) | Delta |
| LC_722 | EPI_ISL_4026613 | B.1.617.2 | 21A (Delta) | Delta |
| LC_723 | EPI_ISL_4026614 | AY.25 | 21A (Delta) | Delta |
| LC_724 | EPI_ISL_4026615 | AY.14 | 21A (Delta) | Delta |
| LC_725 | EPI_ISL_4026616 | AY.25 | 21A (Delta) | Delta |
| LC_727 | EPI_ISL_4026618 | B.1.617.2 | 21A (Delta) | Delta |
| LC_728 | EPI_ISL_4026619 | B.1.617.2 | 21A (Delta) | Delta |
| LC_730 | EPI_ISL_4026620 | AY.3 | 21A (Delta) | Delta |
| LC_731 | EPI_ISL_4026621 | AY.3 | 21A (Delta) | Delta |
| LC_732 | EPI_ISL_4026622 | B.1.617.2 | 21A (Delta) | Delta |
| LC_733 | EPI_ISL_4026623 | AY.3 | 21A (Delta) | Delta |
| LC_735 | EPI_ISL_4026624 | B.1.617.2 | 21A (Delta) | Delta |
| LC_736 | EPI_ISL_4026625 | AY.26 | 21A (Delta) | Delta |
| LC_737 | EPI_ISL_4026626 | AY.25 | 21A (Delta) | Delta |
| LC_739 | EPI_ISL_4026627 | AY.25 | 21A (Delta) | Delta |
| LC_740 | EPI_ISL_4026628 | B.1.617.2 | 21A (Delta) | Delta |
| LC_741 | EPI_ISL_4026629 | B.1.617.2 | 21A (Delta) | Delta |
| LC_742 | EPI_ISL_4026630 | AY.3 | 21A (Delta) | Delta |
| LC_743 | EPI_ISL_4026631 | B.1.617.2 | 21A (Delta) | Delta |
| LC_744 | EPI_ISL_5258008 | B.1.617.2 | 21A (Delta) | Delta |
| LC_745 | EPI_ISL_5258009 | B.1.617.2 | 21A (Delta) | Delta |

| | | | | |
|---|---|---|---|---|
| LC_746 | EPI_ISL_5258010 | AY.25 | 21A (Delta) | Delta |
| LC_747 | EPI_ISL_5258011 | B.1.617.2 | 21A (Delta) | Delta |
| LC_748 | EPI_ISL_5258012 | AY.26 | 21A (Delta) | Delta |
| LC_749 | EPI_ISL_5258013 | AY.20 | 21A (Delta) | Delta |
| LC_750 | EPI_ISL_5258014 | B.1.617.2 | 21A (Delta) | Delta |
| LC_751 | EPI_ISL_5258015 | B.1.617.2 | 21A (Delta) | Delta |
| LC_752 | EPI_ISL_5258016 | B.1.617.2 | 21A (Delta) | Delta |
| LC_753 | EPI_ISL_5258017 | B.1.617.2 | 21A (Delta) | Delta |
| LC_754 | EPI_ISL_5258018 | AY.25 | 21A (Delta) | Delta |
| LC_755 | EPI_ISL_5258019 | AY.26 | 21A (Delta) | Delta |
| LC_756 | EPI_ISL_5258020 | AY.26 | 21A (Delta) | Delta |
| LC_757 | EPI_ISL_5258021 | AY.3 | 21A (Delta) | Delta |
| LC_758 | EPI_ISL_5258026 | AY.3 | 21A (Delta) | Delta |
| LC_759 | EPI_ISL_5258027 | B.1.617.2 | 21A (Delta) | Delta |
| LC_760 | EPI_ISL_5258028 | AY.25 | 21A (Delta) | Delta |
| LC_761 | EPI_ISL_5258029 | AY.34 | 21A (Delta) | Delta |
| LC_762 | EPI_ISL_5258030 | B.1.617.2 | 21A (Delta) | Delta |
| LC_763 | EPI_ISL_5258031 | AY.3 | 21A (Delta) | Delta |
| LC_764 | EPI_ISL_5258032 | B.1.617.2 | 21A (Delta) | Delta |
| LC_765 | EPI_ISL_5258033 | B.1.617.2 | 21A (Delta) | Delta |
| LC_766 | EPI_ISL_5258034 | AY.3 | 21A (Delta) | Delta |
| LC_767 | EPI_ISL_5258035 | B.1.617.2 | 21A (Delta) | Delta |
| LC_768 | EPI_ISL_5258036 | AY.25 | 21A (Delta) | Delta |
| LC_769 | EPI_ISL_5258037 | AY.20 | 21A (Delta) | Delta |
| LC_770 | EPI_ISL_5258038 | B.1.617.2 | 21A (Delta) | Delta |
| LC_771 | EPI_ISL_5258039 | AY.3 | 21A (Delta) | Delta |
| LC_773 | EPI_ISL_5258041 | AY.25 | 21A (Delta) | Delta |
| LC_774 | EPI_ISL_5258042 | B.1.617.2 | 21A (Delta) | Delta |
| LC_775 | EPI_ISL_5258043 | B.1.617.2 | 21A (Delta) | Delta |
| LC_776 | EPI_ISL_5258044 | AY.26 | 21A (Delta) | Delta |
| LC_777 | EPI_ISL_5258045 | B.1.617.2 | 21A (Delta) | Delta |
| LC_778 | EPI_ISL_5258046 | AY.3 | 21A (Delta) | Delta |
| LC_779 | EPI_ISL_5258047 | B.1.617.2 | 21A (Delta) | Delta |
| LC_780 | EPI_ISL_5258048 | B.1.617.2 | 21A (Delta) | Delta |
| LC_781 | EPI_ISL_5258049 | B.1.617.2 | 21A (Delta) | Delta |
| LC_782 | EPI_ISL_5258050 | B.1.617.2 | 21A (Delta) | Delta |
| LC_783 | EPI_ISL_5258051 | AY.3 | 21A (Delta) | Delta |
| LC_784 | EPI_ISL_5258052 | B.1.617.2 | 21A (Delta) | Delta |
| LC_785 | EPI_ISL_5258053 | B.1.617.2 | 21A (Delta) | Delta |

| | | | | |
|---|---|---|---|---|
| LC_786 | EPI_ISL_5258054 | AY.25 | 21A (Delta) | Delta |
| LC_787 | EPI_ISL_5258055 | B.1.617.2 | 21A (Delta) | Delta |
| LC_788 | EPI_ISL_5258056 | B.1.617.2 | 21A (Delta) | Delta |
| LC_789 | EPI_ISL_5258057 | AY.3 | 21A (Delta) | Delta |
| LC_791 | EPI_ISL_5258059 | B.1.617.2 | 21A (Delta) | Delta |
| LC_792 | EPI_ISL_5258060 | B.1.617.2 | 21A (Delta) | Delta |
| LC_793 | EPI_ISL_5258061 | B.1.617.2 | 21A (Delta) | Delta |

Based on the lineage of SARS-CoV-2 identified by WGS, children were grouped based on whether their COVID-19 infection was caused by a VOC, as well as the specific VOC lineage, using CDC definitions as of August 19$^{th}$, 2021.[2]

**eTable 2: Medical conditions with high-risk of COVID-19 complications**

| Medical Condition | Highest Level of Evidence |
|---|---|
| Bronchiectasis | Meta-Analysis and/or systematic review |
| Bronchopulmonary dysplasia | |
| Pulmonary hypertension and pulmonary embolism | |
| Cancer | |
| Cerebrovascular disease | |
| Chronic kidney disease | |
| Chronic liver disease | |
| COPD | |
| Diabetes mellitus, type 1 | |
| Diabetes mellitus, type 2 | |
| Heart conditions | |
| Interstitial lung disease | |
| Smoking, current and former | |
| Tuberculosis | |
| Obesity* | |
| Pregnancy and Recent Pregnancy | |
| Mental health disorders | |
| Down syndrome | Observational study |
| HIV | |
| Neurologic conditions | |
| Overweight* | |
| Sickle cell disease | |
| Solid organ or blood stem cell transplantation | |
| Substance use disorders | |
| Use of corticosteroids or other immunosuppressive medications | |

Children in this study were classified as high risk for COVID-19 complications if they had an underlying condition for which there are high-quality and reproducible data (i.e., meta-analysis, systematic review, or observational study; excluding small studies, case reports/series, or conflicting evidence) based on CDC classification as of August 31st, 2021.[3]

*Obesity identified as body mass index (BMI) ≥30 kg/m$^2$ (or ≥95%ile), and overweight defined as BMI 25-30 (or 85-95%ile). If height was unknown and BMI was undetermined, patient was identified as high risk if weight > 95%ile.

**eTable 3: COVID-19 World Health Organization Clinical Progression Scale[4]**

| Patient State | Descriptor | Score |
|---|---|---|
| Uninfected | Uninfected; no viral RNA detected | 0 |
| Ambulatory mild disease | Asymptomatic; viral RNA detected | 1 |
| | Symptomatic; independent | 2 |
| | Symptomatic; assistance needed | 3 |
| Hospitalized: moderate disease | Hospitalized; no oxygen therapy* | 4 |
| | Hospitalized; oxygen by mask or nasal cannula | 5 |
| Hospitalized: severe disease | Hospitalized; oxygen by non-invasive ventilation or high-flow nasal cannula | 6 |
| | Intubation and mechanical ventilation, $pO_2/FiO_2 \geq 150$ or $SpO_2/FiO_2 \geq 200$ | 7 |
| | Mechanical ventilation $pO_2/FiO_2 < 150$ ($SpO_2/FiO_2 < 200$) or vasopressors | 8 |
| | Mechanical ventilation $pO_2/FiO_2$ and vasopressors, dialysis, or extracorporeal membrane oxygenation | 9 |
| Dead | Dead | 10 |

$pO_2$: partial pressure of oxygen. $FiO_2$: fraction of inspired oxygen. $SpO_2$: oxygen saturation.

*If hospitalized for reasons other than COVID-19, status recorded as for ambulatory patient.

**eTable 4: Sensitivity analysis^ assessing association between COVID-19 severity and SARS-CoV-2 variants of concern correcting for potential impact of COVID-19 vaccine and monoclonal antibodies**

| | **Marker of COVID-19 Severity: Odds Ratio (95% Confidence Interval; *p* Value)** | | | | |
|---|---|---|---|---|---|
| | **Hospitalized for COVID-19** | **COVID-19 Pharmacologic Treatment** | **Respiratory Support** | **Intensive Care Unit Admission** | **WHO Clinical Progression Scale Score ≥6** |
| Age | 1.02 (0.96-1.09; 0.5) | **1.2 (1.1-1.4; <0.001)*** | **1.2 (1.1-1.3; 0.003)*** | **1.2 (1.1-1.3; 0.003)*** | **1.2 (1.1-1.4; 0.003)*** |
| Race (Black) | 1.03 (0.36-2.7; >0.9) | 0.60 (0.15-2.1; 0.4) | 0.53 (0.11-2.1; 0.4) | 0.61 (0.12-2.4; 0.5) | 0.7 (0.12-2.8; 0.6) |
| Ethnicity (Hispanic) | 1.3 (0.56-2.8; 0.6) | 0.43 (0.12-1.3; 0.2) | 0.44 (0.11-1.5; 0.2) | 0.45 (0.11-1.6; 0.2) | 0.43 (0.08-1.7; 0.3) |
| High-risk condition for COVID-19 complications | **5.1 (2.4-12.0, <0.001)*** | - | - | - | - |
| Pediatric COVID-19 community incidence | 1.0 (1.0-1.0; 0.6) | 1.0 (1.0-1.0; 0.2) | 1.0 (1.0-1.0; 0.8) | 1.0 (1.0-1.0; 0.2) | 1.0 (1.0-1.0; 0.7) |
| SARS-CoV-2 lineage | | | | | |
| *Non-VOC* | ref | ref | ref | Ref | Ref |
| *Alpha VOC* | 2.8 (0.95-8.4; 0.06) | 2.1 (0.5-9.4; 0.3) | 2.1 (0.34-12.9; 0.4) | 2.2 (0.43-11.9; 0.3) | 2.8 (0.4-24.3; 0.3) |
| *Gamma VOC* | **6.5 (1.9-22.7; 0.003)*** | 4.7 (0.97-23.2; 0.05) | **10.6 (2.2-65.0; 0.005)*** | 4.3 (0.72-26.1; 0.1) | **11.2 (1.8-98.1; 0.013)*** |
| *Delta VOC* | 2.5 (0.80-8.1; 0.12) | 2.3 (0.53-10.8; 0.3) | 3.1 (0.57-19.9; 0.2) | 2.2 (0.43-12.7; 0.3) | 3.3 (0.46-31.0; 0.2) |

VOC- variant of concern. y- years. m- month. ref- reference group for VOC odds ratio calculations. N/A- not applicable. High-risk conditions were excluded from the model for some outcomes because having a high-risk condition was mutually inclusive with the outcome of interest.

^A small number of patients were fully vaccinated (n=2; one each in the gamma and delta VOC groups) at time of their breakthrough infection or had received SARS-CoV-2 monoclonal antibodies (n=3; all in the alpha VOC group) as an outpatient early in infection to prevent subsequent morbidity. A sensitivity analysis was performed whereby we repeated the models reported in Table 1, presuming that these five patients would have experienced each severity marker had they not received these therapies. *Bolded values indicate statistical significance (*p*<0.05).

**Supplemental References**

1. Grubaugh ND, Gangavarapu K, Quick J, et al. An amplicon-based sequencing framework for accurately measuring intrahost virus diversity using PrimalSeq and iVar. *Genome biology*. Jan 8 2019;20(1):8. doi:10.1186/s13059-018-1618-7
2. Centers for Disease Control and Prevention; SARS-CoV-2 Variant Classifications and Definitions. Accessed August 31st, 2021, https://www.cdc.gov/coronavirus/2019-ncov/variants/variant-info.html
3. Centers for Disease Control and Prevention; Science Brief: Evidence Used to Update the List of Underlying Medical Conditions Associated with Higher Risk for Severe COVID-19. Accessed August 31st, 2021, https://www.cdc.gov/coronavirus/2019-ncov/science/science-briefs/underlying-evidence-table.html
4. WHO Working Group on the Clinical Characterisation and Management of COVID-19 infection; A minimal common outcome measure set for COVID-19 clinical research. *Lancet Infect Dis*. Aug 2020;20(8):e192-e197. doi:10.1016/s1473-3099(20)30483-7
